# Prevalence of anxiety and depression symptoms under population-level alcohol reduction to low-risk drinking: A parametric g-computation analysis in a U.S. nationwide cohort

**DOI:** 10.64898/2026.07.31.26359417

**Authors:** Jenna Sanborn, Denis Nash, McKaylee Robertson, Angela Parcesepe, Zachary Shahn

## Abstract

**Background:** Alcohol consumption is a modifiable risk factor linked to anxiety and depression, yet the population-level impact of reducing alcohol use on the prevalence of anxiety and depression symptoms remains unclear. Prior studies are limited by methodological challenges including health-related selection into abstinence, time-varying confounding and reverse causation.

**Methods:** We used data from 2,813 participants without documented underlying health conditions in a U.S. national longitudinal cohort (September 2021-December 2023) and applied the parametric g-formula within a target trial emulation framework to estimate the effect of a hypothetical intervention reducing alcohol consumption to low-risk levels on the prevalence of moderate-to-severe anxiety (GAD-7 ≥10) and depression (PHQ-8 ≥10) symptoms. The intervention set moderate- and high-risk drinking to low-risk at each follow-up, while leaving abstinent and low-risk drinking unchanged. Models adjusted for time-varying and baseline covariates, with uncertainty estimated using bootstrap resampling.

**Results:** Under the natural course, the estimated prevalence of moderate-to-severe depressive symptoms was 12.67% (95% CI: 12.17, 13.23), compared to 12.77% (95% CI: 12.26, 13.29) under the intervention (risk difference: 0.10 percentage points; 95% CI: -0.22, 0.50). For anxiety symptoms, prevalence was 10.40% (95% CI: 8.89, 11.89) under the natural course and 10.25% (95% CI: 8.76, 11.98) under the intervention (risk difference: -0.15 percentage points; 95% CI: - 0.68, 0.35).

**Conclusions:** Under a hypothetical population-wide intervention reducing alcohol consumption to low-risk levels, differences in the prevalence of moderate or severe anxiety and depression symptoms were near null. These findings suggest that reducing alcohol use alone may be insufficient to meaningfully shift the population-wide burden of common mental health disorders.

## Introduction

Anxiety and depression are prevalent public health concerns in the United States (U.S.), affecting approximately 23% of adults and consistently ranking among leading causes of global disability.^1–3^ Alcohol consumption is also widespread, with more than 174 million adults reporting alcohol use.^4^ Patterns of heavy drinking are common: approximately 57 million adults (21.7%) report binge drinking in the past month and 28 million (10.9%) meet criteria for alcohol use disorder.^4,5^ Alcohol use disorder is strongly linked to depression and anxiety, with systematic reviews supporting a bidirectional causal relationship between alcohol use disorder and major depression and anxiety disorders.^6,7^ Those with alcohol use disorder are more than twice as likely to have major depression (pooled OR 2.42, 95% CI 2.22-2.64) or an anxiety disorder (pooled OR 2.11, 95% CI 2.03-2.19) compared to those without.^8,9^

An extensive body of observational research has examined associations between varying levels of alcohol consumption and anxiety and depression. Most community-based cross-sectional and longitudinal studies report that heavy or hazardous drinking is associated with higher odds of anxiety and depression compared to abstinence.^10–15^ Findings regarding moderate alcohol consumption are less consistent, with many reporting a J- or U-shaped relationship in which moderate drinkers have better mental health outcomes than abstainers and heavy drinkers.^11,12,16–23^ However, this literature is subject to several documented methodological challenges, including health-related selection into abstinence (sick-quitter bias), reverse causation, and time-varying confounding, which can lead to biased estimates.^6,24–27^ These challenges are compounded by the non-absorptive and cyclical nature of anxiety and depression symptoms, with individuals transitioning into and out of symptomatic periods over time, such that mental health may both influence and be influenced by alcohol consumption. Thus, approaches that leverage causal inference methods that address these limitations are urgently needed.^28,29^

Because alcohol consumption is a modifiable behavioral target, estimating how the prevalence of anxiety and depression might change under realistic reductions in alcohol consumption may provide policy-relevant evidence on the potential impact of alcohol reduction strategies and help inform public health strategies aimed at reducing the burden of common mental health disorders. To the best of our knowledge, no empirical evidence exists regarding how population-level reductions might influence the prevalence of anxiety and depression symptoms. The parametric g-formula, also known as g-computation, is a model-based standardization approach that estimates population-level outcomes under hypothetical exposure interventions.^26,30^ In doing so, it enables estimation of the outcome risk that would have been observed if exposure patterns had differed from those actually experienced in the cohort.^31^ By modeling longitudinal relationships among exposures, outcomes, and other time-varying covariates, g-computation adjusts for confounders that change over time and may be affected by prior exposure, helping to reduce bias from reverse causation and time-dependent confounding.

In this study, we used longitudinal data from a nationwide cohort of U.S. adults and applied the parametric g-formula to estimate how the prevalence of moderate to severe anxiety and depression symptoms would change under a hypothetical population-level reduction in alcohol consumption to low-risk levels among individuals reporting moderate or high-risk drinking. To reduce bias related to sick-quitter bias, we restricted the analysis to adults without documented health conditions that might lead individuals to reduce or stop drinking, consistent with prior findings from this cohort showing that the protective association between moderate drinking and anxiety and depression disappeared after excluding participants with underlying conditions.^32^ By estimating mental health outcomes under a realistic alcohol reduction scenario, this study provides evidence relevant to potential mental health benefits of alcohol reduction strategies at the population level.

## Methods

### Study Population and Target Trial Specification

The CHASING COVID Cohort is a nationwide prospective study of 5,798 adults aged 18 years or older recruited online between March-August 2020. Participants resided in all 50 U.S. states, Washington D.C., Puerto Rico, and Guam, resulting in a geographically and socio-demographically diverse population. Assessments were administered approximately quarterly through December 2023. Recruitment procedures and follow-up protocols have been described previously,^33^ and study materials are publicly available.^34^ The assessments collected repeated information on health-related experiences, behaviors, and socio-demographic characteristics.

For this analysis, a target trial was emulated to estimate the effect of sustained alcohol consumption patterns on prevalence of moderate to severe anxiety and depression symptoms (Table 1). Eligibility was defined at the September 2021 assessment, which served as the analytic baseline and the start of follow-up. This timepoint corresponded to the post-vaccine period when the acute phase of the pandemic had stabilized and major pandemic-related disruptions had largely subsided. Participants were followed from September 2021 through December 2023. Alcohol consumption was assessed at five assessments during follow-up (September 2021, March 2022, October 2022, April 2023, and September 2023). Symptoms of anxiety and depression were assessed at ten timepoints, with the primary outcome defined using measurements from the end of follow-up (December 2023). Time-varying covariates included repeated measures of anxiety and depression symptoms, mental health diagnoses and treatment, as well as indicators of health status, socioeconomic conditions, and health behaviors.

**Table 1.** Specification of the target trial and its emulation.

| Table 1. Specification of the target trial and its emulation |  |  |
| --- | --- | --- |
| Component | Target Trial (Ideal RCT) | Emulation in This Study |
| Eligibility | Adults without underlying health conditions | Adults without underlying health conditions at September 2021 (baseline) |
| Treatment strategies | Assign participants to maintain usual drinking vs reduce to low-risk levels | Observed alcohol use; intervention simulated by setting alcohol consumption to low-risk levels at each timepoint |
| Treatment assignment | Assign participants to maintain usual drinking versus reduce to low-risk levels | Observed alcohol use; intervention simulated by setting moderate- and high-risk alcohol consumption to low-risk levels at each timepoint, while leaving abstinent and low-risk drinking unchanged |
| Follow-up | Fixed follow-up period | September 2021 to December 2023 |
| Outcome | PHQ-8 and GAD-7 scores at end of follow-up | PHQ-8 and GAD-7 scores at end of follow-up; moderate-to-severe symptoms defined as PHQ-8 ≥10 or GAD-7 ≥10 |
| Causal contrast | Difference in risk under each strategy | Difference in estimated risk under intervention vs natural course |
| Confounding control | Randomization | Adjustment for measured time-varying confounders using the parametric g-formula |
| Identifiability conditions | Exchangeability by randomization, positivity by design, consistency, well-defined treatment strategies, and no interference between units | Causal interpretation requires no unmeasured confounding conditional on measured covariate history; no positivity violations; consistency between observed outcomes and potential outcomes under the corresponding alcohol-use histories; a sufficiently well-defined intervention reducing moderate- or high-risk AUDIT-C drinking to low-risk drinking; no interference between participants; and correct specification of the parametric models used in the g-formula |

### Analytic Sample

Of the 5,798 CHASING COVID cohort participants, 4,684 completed the September 2021 assessment, which served as the analytic baseline for this study. Among those completing the baseline assessment, we excluded individuals missing sex data (n = 11) and those reporting chronic conditions or poor general health at baseline (n = 1,860). Chronic conditions were defined based on self-reported diagnoses (including diabetes, high blood pressure, chronic lung disease or COPD, heart attack, angina, kidney disease, immunocompromised condition, HIV, or current asthma) at enrollment. These individuals were excluded because poor health or chronic conditions may plausibly lead individuals to reduce or stop drinking, which could introduce bias due to health-related changes in alcohol consumption, which we were unable to fully account for given limited measurement of these processes. Results should therefore be interpreted to be among adults without documented health conditions.

The final analytic sample included 2,813 participants (Figure 1). Baseline characteristics of the analytic sample compared with excluded participants are presented in Supplementary Table S1. Participants included in the analytic sample generally exhibited lower prevalence of depression and anxiety symptoms and differed modestly in alcohol use patterns compared with those excluded.

**Figure 1.**
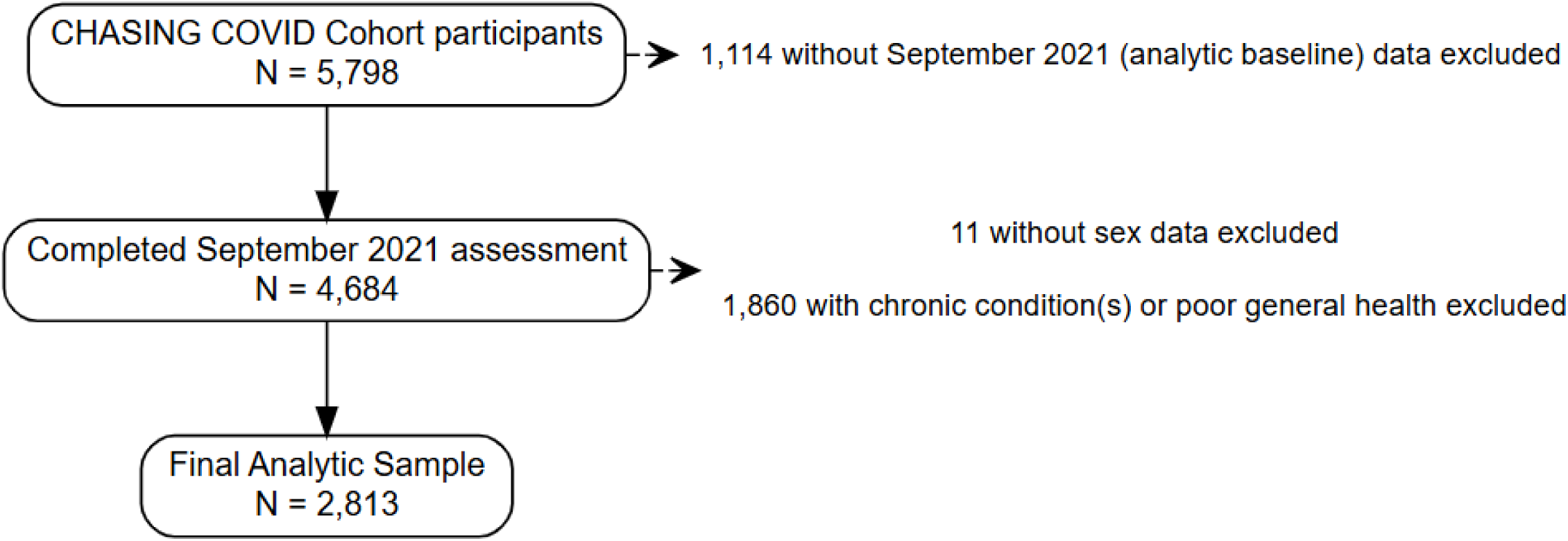
Study Flow Diagram.

### Research Ethics

The study protocol was approved by the Institutional Review Board at the City University of New York (CUNY). Participant consent was obtained at baseline and at periodic follow-up assessments. Participants could voluntarily discontinue participation at any time.

### Alcohol consumption (exposure)

Alcohol consumption was assessed using the 3-item Alcohol Use Disorders Identification Test-Consumption (AUDIT-C), a validated screening tool with high sensitivity and specificity for identifying hazardous drinking.^35^ The scale sums three items on drinking frequency, typical quantity, and frequency of binge drinking, with scores ranging from 0 (no use) to 12 where higher scores indicate a higher likelihood of alcohol misuse.^36^ We categorized consumption into abstinence, low risk, moderate risk, and high/severe risk groups, using sex-specific cutoff scores based on guidelines adapted from the U.S. Department of Veterans Affairs: abstinence (score = 0 for both sexes), low risk (scores 1-2 for females, 1-3 for males), moderate risk (scores 3-5 for females, 4-5 for males), and high/severe risk (score ≥6 for both sexes)..^37^

### Anxiety and depression symptoms (outcomes)

Symptoms of anxiety and depression were measured using the Generalized Anxiety Disorder-7 (GAD-7; score range 0-21) and Patient Health Questionnaire-8 (PHQ-8; score range 0-24). Scores ≥10 on either scale indicate moderate to severe symptoms. Both instruments are validated and widely used in research and clinical settings.^38–42^

### Covariates

Supplemental Table S2 provides detailed descriptions of all covariates included in the analysis. A full list of covariates and the specific forms for each used in the models can be found in Table 3.

Time-varying covariates included measures of mental health (moderate-to-severe anxiety or depression symptoms, anxiety or depression diagnosis), mental health treatment (receipt of psychiatric medication and receipt of psychotherapy), health status (disability, poor physical-health days, poor mental health days, and general health status), socioeconomic factors (housing instability, food insecurity, employment status, and participation in government food-support programs), health behaviors (minutes of physical activity per week, drug use, and SARS-CoV-2 vaccination status), and healthcare access measures, and chronic conditions diagnosed during follow-up. Covariate selection was determined a priori based on prior literature, and directed acyclic graphs identifying variables plausibly associated with both alcohol exposure and anxiety and depression. To avoid adjustment for covariates impacted by exposure, at each timepoint at which the exposure was assessed, we adjusted for covariate history summarized through the immediately preceding assessment. However, in the instances when a covariate was not measured at the prior wave, the value from the concurrent assessment was used.

We included time-fixed covariates measured prior to September 2021 including age, gender, race/ethnicity, educational attainment, income, body mass index, daily smoking status, household size, residential area type, income loss, social engagement level, relationship status, alcohol or drug-use recovery status, neighborhood community, life satisfaction, and spirituality or religion.

To account for participants’ behavioral and mental health histories, we additionally included pre-baseline summary measures calculated from earlier cohort assessments. These included the mean AUDIT-C score, any high-risk drinking, mean GAD-7 score, mean PHQ-8 score, mean weekly physical activity, any food insecurity, and mean housing insecurity score, prior to baseline. Given that housing insecurity was assessed on a 5-point Likert scale (ranging from “never” to “always” worried about paying rent or mortgage), the average score reflects typical level of financial stress related to housing.

### Statistical analysis

We sought to estimate the effect of sustained alcohol reduction on the prevalence of moderate-to-severe symptoms of depression and anxiety under a target trial emulation framework. We compared outcomes under two scenarios: the natural course of alcohol use (the level it would take absent any intervention) and a hypothetical intervention that capped alcohol consumption at low-risk levels at each follow-up assessment. This intervention was specified to represent a more plausible population-level contrast than complete abstinence. The primary estimand was the difference in risk of moderate-to-severe symptoms under the intervention versus the natural course.

We implemented the parametric g-formula using Monte Carlo simulation. The g-formula was used to simulate end-of-follow-up outcomes under both scenarios, from which we estimated risks and risk differences for moderate-to-severe depression (PHQ-8 ≥10) and anxiety (GAD-7 ≥10). We focused on prevalence rather than differences in continuous symptom scores because the proportion of individuals experiencing clinically meaningful symptoms more directly reflects population burden. Ninety-five percent confidence intervals were obtained using a nonparametric bootstrap. Additional details on model specification, simulation procedures, and diagnostics are provided in the Supplement. A diagram summarizing the steps used to carry out the g-computation analysis using a continuous end-of-follow-up outcome can be found in Figure 2.

**Figure 2.**
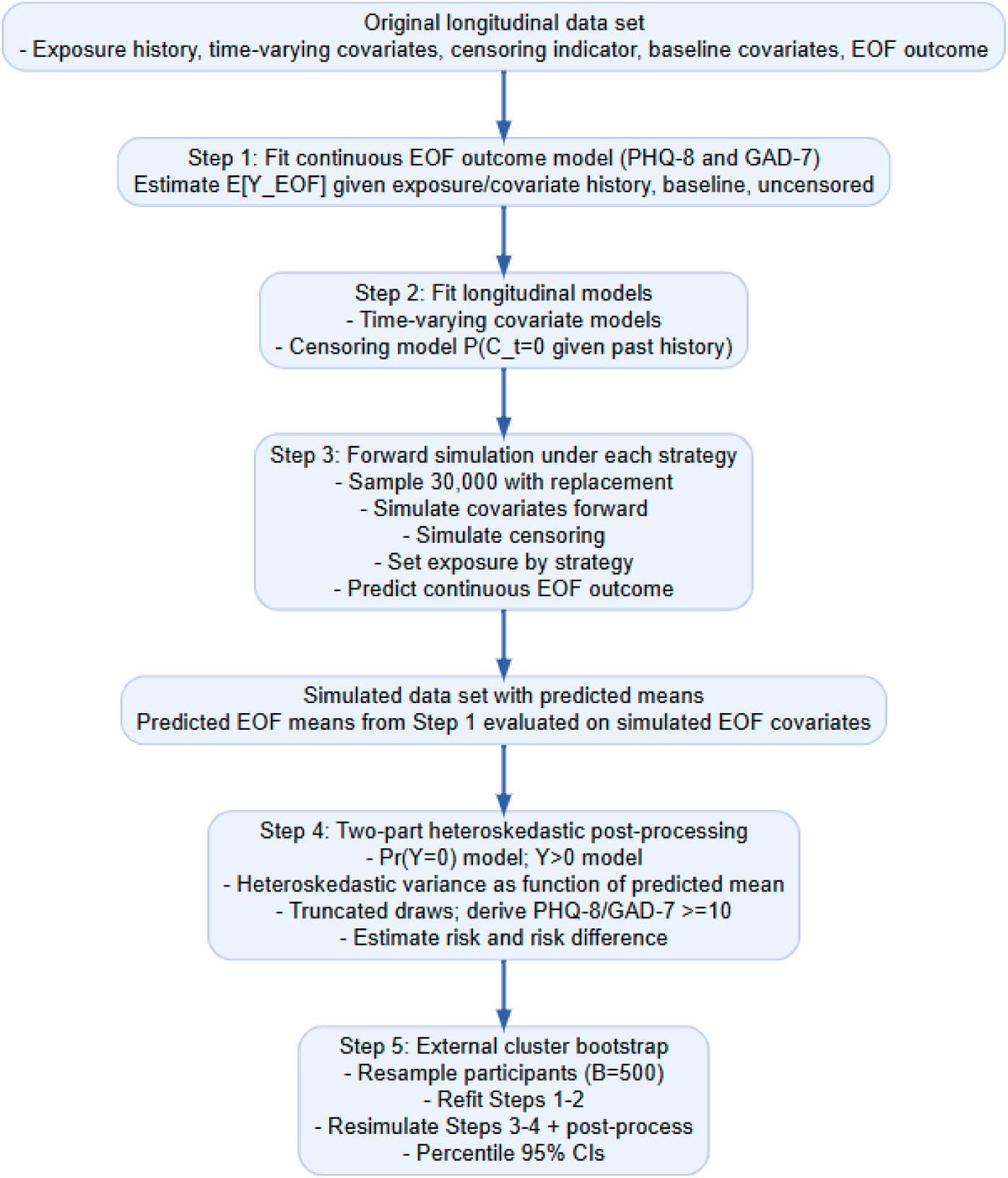
Schematic of the g-computation procedure.

We conducted sensitivity analyses varying the timing of time-varying covariates (using concurrent rather than lagged values to assess sensitivity to temporal ordering) and excluding participants in recovery from alcohol or drug use at baseline, to evaluate potential bias from recovery-related changes in drinking behavior (details in Supplement).

Analyses were conducted in R (version 4.5.2) using the gfoRmula package.

## Results

Among the 2,813 participants included in the analytic sample, 27.6% were abstinent from alcohol, 37.8% were low-risk drinkers, 25.3% were moderate-risk drinkers, and 9.3% were high/severe-risk drinkers in September 2021 (baseline) (Table 2). The prevalence of moderate/severe anxiety and depression symptoms was highest among participants reporting high/severe-risk drinking (24.4% and 27.1%, respectively) and lowest among those reporting moderate-risk drinking (14.6% and 16.3%), with slightly higher prevalence observed among low-risk (14.7% and 17.6%) and abstinent participants (19.7% and 21.0%). Sociodemographic, behavioral, and health-related characteristics varied across alcohol consumption categories (Table 2).

**Table 2.**
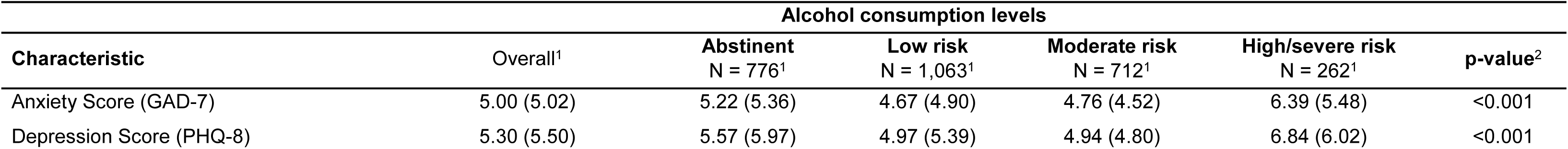
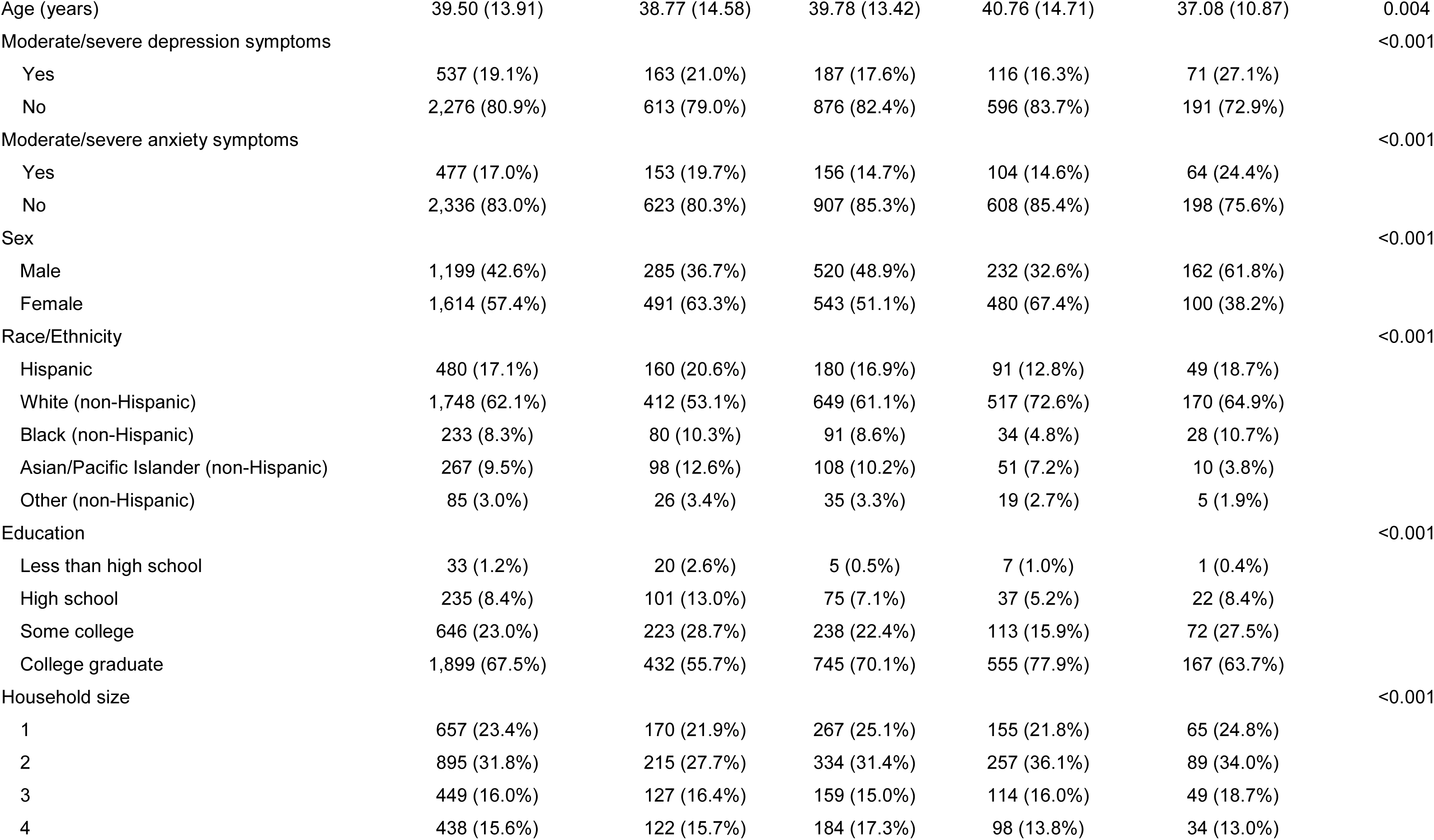
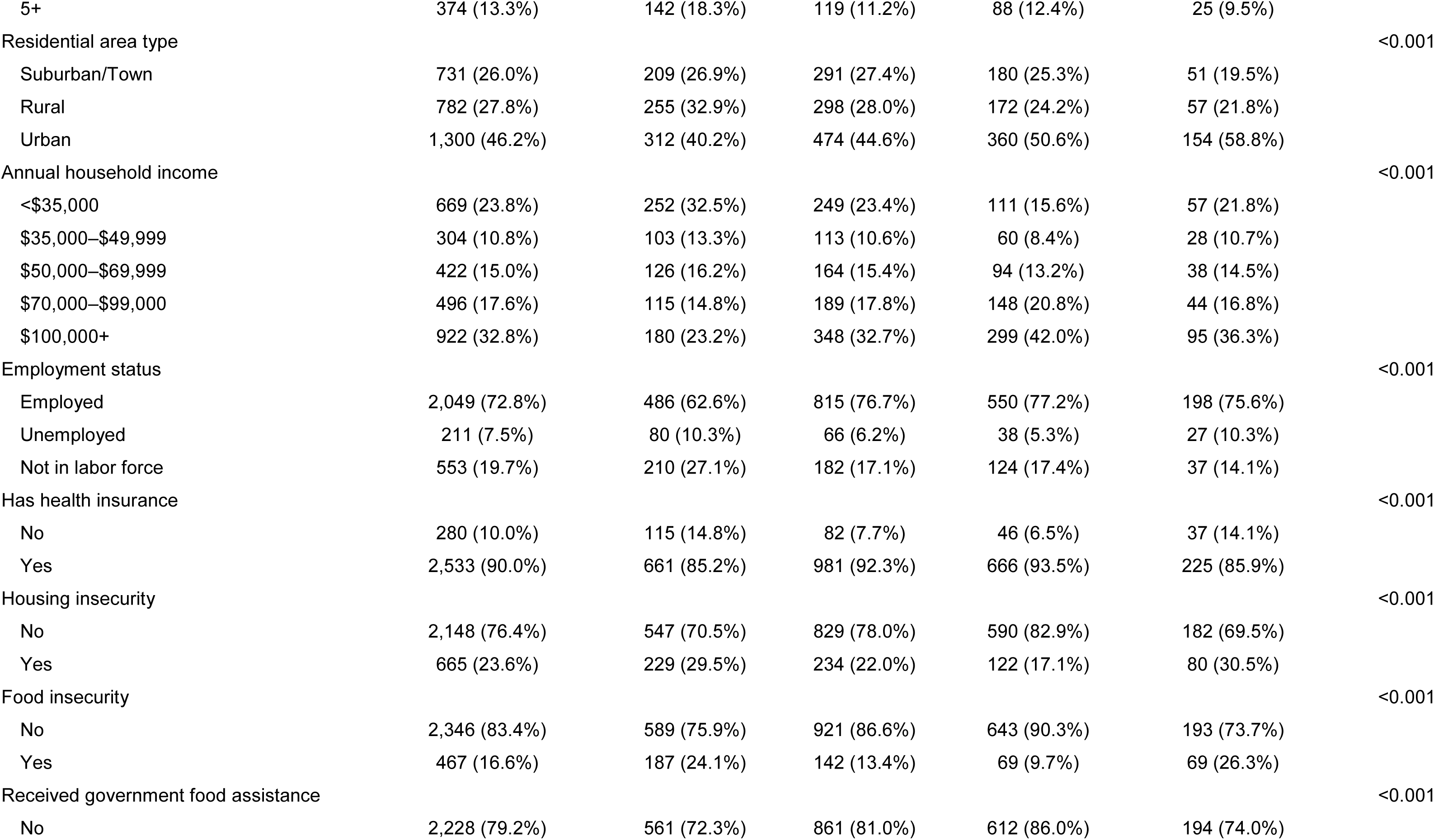
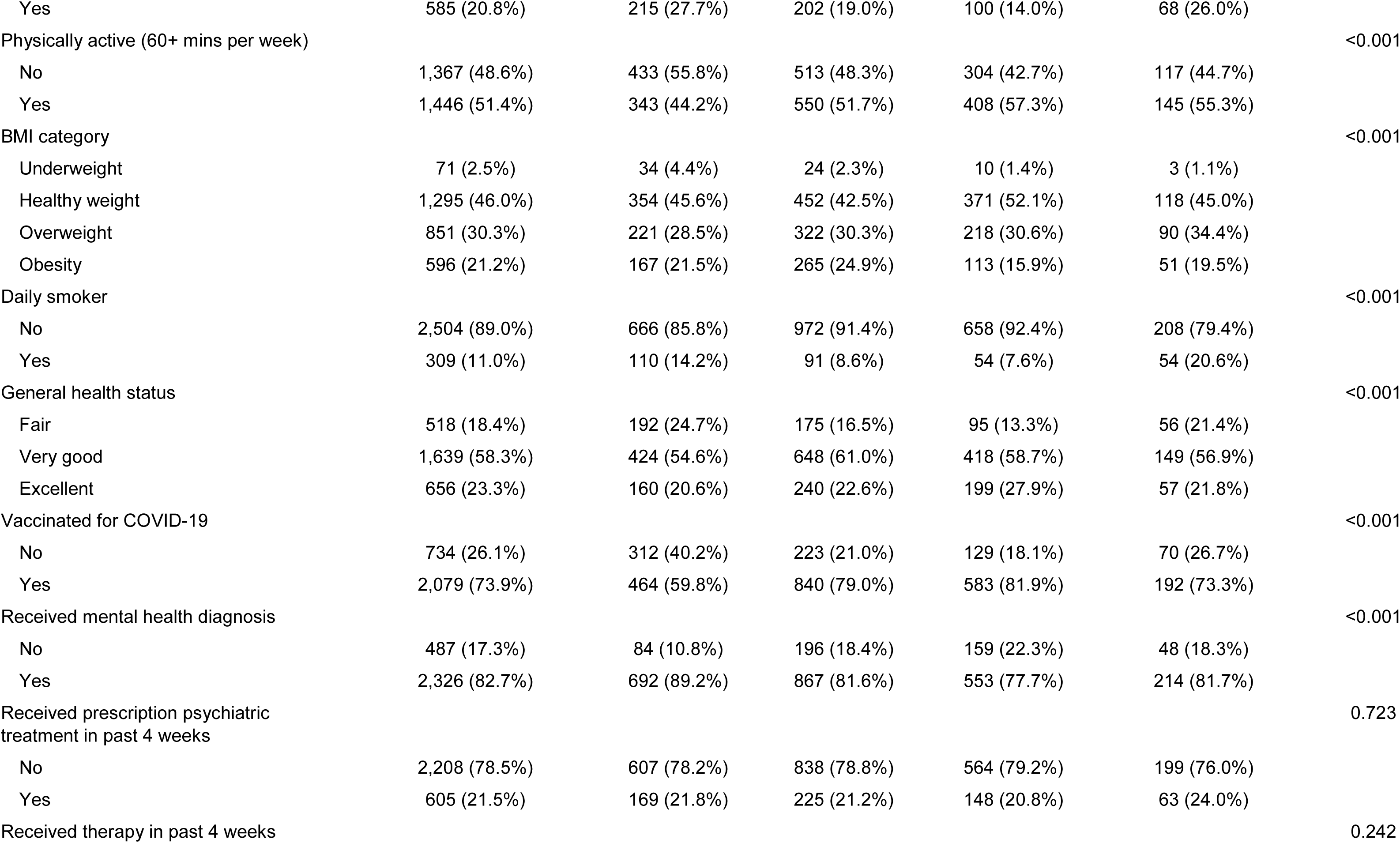
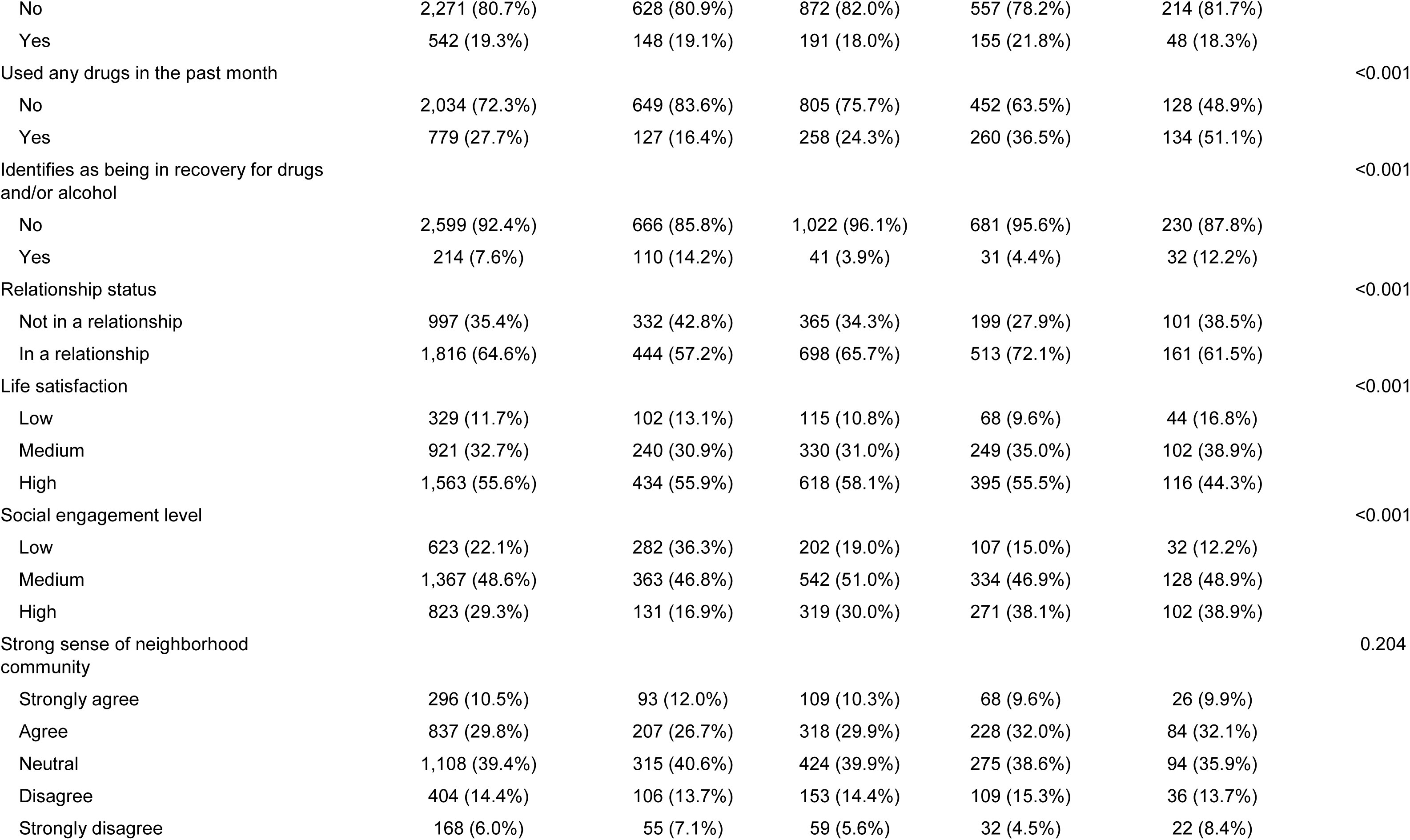
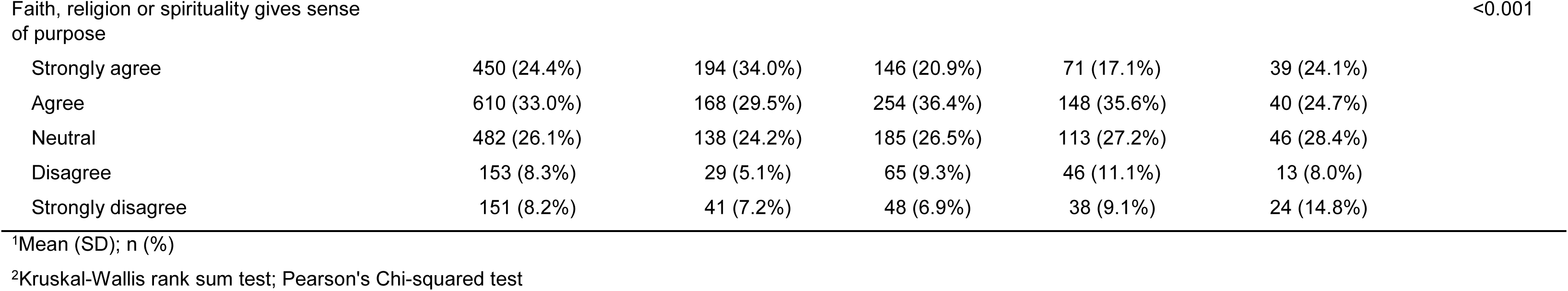
Characteristics of the sample by exposure status at analytic baseline, CHASING COVID Cohort, September 2021 (N=2,813)

| Alcohol consumption levels |  |  |  |  |  |  |
| --- | --- | --- | --- | --- | --- | --- |
| Characteristic | Overall<sup>1</sup> | Abstinent<br>N = 776<sup>1</sup> | Low risk<br>N = 1,063<sup>1</sup> | Moderate risk<br>N = 712<sup>1</sup> | High/severe risk<br>N = 262<sup>1</sup> | p-value<sup>2</sup> |
| Anxiety Score (GAD-7) | 5.00 (5.02) | 5.22 (5.36) | 4.67 (4.90) | 4.76 (4.52) | 6.39 (5.48) | <0.001 |
| Depression Score (PHQ-8) | 5.30 (5.50) | 5.57 (5.97) | 4.97 (5.39) | 4.94 (4.80) | 6.84 (6.02) | <0.001 |
| Age (years) | 39.50 (13.91) | 38.77 (14.58) | 39.78 (13.42) | 40.76 (14.71) | 37.08 (10.87) | 0.004 |
| Moderate/severe depression symptoms |  |  |  |  |  | <0.001 |
| Yes | 537 (19.1%) | 163 (21.0%) | 187 (17.6%) | 116 (16.3%) | 71 (27.1%) |  |
| No | 2,276 (80.9%) | 613 (79.0%) | 876 (82.4%) | 596 (83.7%) | 191 (72.9%) |  |
| Moderate/severe anxiety symptoms |  |  |  |  |  | <0.001 |
| Yes | 477 (17.0%) | 153 (19.7%) | 156 (14.7%) | 104 (14.6%) | 64 (24.4%) |  |
| No | 2,336 (83.0%) | 623 (80.3%) | 907 (85.3%) | 608 (85.4%) | 198 (75.6%) |  |
| Sex |  |  |  |  |  | <0.001 |
| Male | 1,199 (42.6%) | 285 (36.7%) | 520 (48.9%) | 232 (32.6%) | 162 (61.8%) |  |
| Female | 1,614 (57.4%) | 491 (63.3%) | 543 (51.1%) | 480 (67.4%) | 100 (38.2%) |  |
| Race/Ethnicity |  |  |  |  |  | <0.001 |
| Hispanic | 480 (17.1%) | 160 (20.6%) | 180 (16.9%) | 91 (12.8%) | 49 (18.7%) |  |
| White (non-Hispanic) | 1,748 (62.1%) | 412 (53.1%) | 649 (61.1%) | 517 (72.6%) | 170 (64.9%) |  |
| Black (non-Hispanic) | 233 (8.3%) | 80 (10.3%) | 91 (8.6%) | 34 (4.8%) | 28 (10.7%) |  |
| Asian/Pacific Islander (non-Hispanic) | 267 (9.5%) | 98 (12.6%) | 108 (10.2%) | 51 (7.2%) | 10 (3.8%) |  |
| Other (non-Hispanic) | 85 (3.0%) | 26 (3.4%) | 35 (3.3%) | 19 (2.7%) | 5 (1.9%) |  |
| Education |  |  |  |  |  | <0.001 |
| Less than high school | 33 (1.2%) | 20 (2.6%) | 5 (0.5%) | 7 (1.0%) | 1 (0.4%) |  |
| High school | 235 (8.4%) | 101 (13.0%) | 75 (7.1%) | 37 (5.2%) | 22 (8.4%) |  |
| Some college | 646 (23.0%) | 223 (28.7%) | 238 (22.4%) | 113 (15.9%) | 72 (27.5%) |  |
| College graduate | 1,899 (67.5%) | 432 (55.7%) | 745 (70.1%) | 555 (77.9%) | 167 (63.7%) |  |
| Household size |  |  |  |  |  | <0.001 |
| 1 | 657 (23.4%) | 170 (21.9%) | 267 (25.1%) | 155 (21.8%) | 65 (24.8%) |  |
| 2 | 895 (31.8%) | 215 (27.7%) | 334 (31.4%) | 257 (36.1%) | 89 (34.0%) |  |
| 3 | 449 (16.0%) | 127 (16.4%) | 159 (15.0%) | 114 (16.0%) | 49 (18.7%) |  |
| 4 | 438 (15.6%) | 122 (15.7%) | 184 (17.3%) | 98 (13.8%) | 34 (13.0%) |  |
| 5+ | 374 (13.3%) | 142 (18.3%) | 119 (11.2%) | 88 (12.4%) | 25 (9.5%) |  |
| Residential area type |  |  |  |  |  | <0.001 |
| Suburban/Town | 731 (26.0%) | 209 (26.9%) | 291 (27.4%) | 180 (25.3%) | 51 (19.5%) |  |
| Rural | 782 (27.8%) | 255 (32.9%) | 298 (28.0%) | 172 (24.2%) | 57 (21.8%) |  |
| Urban | 1,300 (46.2%) | 312 (40.2%) | 474 (44.6%) | 360 (50.6%) | 154 (58.8%) |  |
| Annual household income |  |  |  |  |  | <0.001 |
| <\$35,000 | 669 (23.8%) | 252 (32.5%) | 249 (23.4%) | 111 (15.6%) | 57 (21.8%) | |
| \$35,000–\$49,999 | 304 (10.8%) | 103 (13.3%) | 113 (10.6%) | 60 (8.4%) | 28 (10.7%) | |
| \$50,000–\$69,999 | 422 (15.0%) | 126 (16.2%) | 164 (15.4%) | 94 (13.2%) | 38 (14.5%) | |
| \$70,000–\$99,000 | 496 (17.6%) | 115 (14.8%) | 189 (17.8%) | 148 (20.8%) | 44 (16.8%) | |
| \$100,000+ | 922 (32.8%) | 180 (23.2%) | 348 (32.7%) | 299 (42.0%) | 95 (36.3%) | |
| Employment status |  |  |  |  |  | <0.001 |
| Employed | 2,049 (72.8%) | 486 (62.6%) | 815 (76.7%) | 550 (77.2%) | 198 (75.6%) |  |
| Unemployed | 211 (7.5%) | 80 (10.3%) | 66 (6.2%) | 38 (5.3%) | 27 (10.3%) |  |
| Not in labor force | 553 (19.7%) | 210 (27.1%) | 182 (17.1%) | 124 (17.4%) | 37 (14.1%) |  |
| Has health insurance |  |  |  |  |  | <0.001 |
| No | 280 (10.0%) | 115 (14.8%) | 82 (7.7%) | 46 (6.5%) | 37 (14.1%) |  |
| Yes | 2,533 (90.0%) | 661 (85.2%) | 981 (92.3%) | 666 (93.5%) | 225 (85.9%) |  |
| Housing insecurity |  |  |  |  |  | <0.001 |
| No | 2,148 (76.4%) | 547 (70.5%) | 829 (78.0%) | 590 (82.9%) | 182 (69.5%) |  |
| Yes | 665 (23.6%) | 229 (29.5%) | 234 (22.0%) | 122 (17.1%) | 80 (30.5%) |  |
| Food insecurity |  |  |  |  |  | <0.001 |
| No | 2,346 (83.4%) | 589 (75.9%) | 921 (86.6%) | 643 (90.3%) | 193 (73.7%) |  |
| Yes | 467 (16.6%) | 187 (24.1%) | 142 (13.4%) | 69 (9.7%) | 69 (26.3%) |  |
| Received government food assistance |  |  |  |  |  | <0.001 |
| No | 2,228 (79.2%) | 561 (72.3%) | 861 (81.0%) | 612 (86.0%) | 194 (74.0%) |  |
| Yes | 585 (20.8%) | 215 (27.7%) | 202 (19.0%) | 100 (14.0%) | 68 (26.0%) |  |
| Physically active (60+ mins per week) |  |  |  |  |  | <0.001 |
| No | 1,367 (48.6%) | 433 (55.8%) | 513 (48.3%) | 304 (42.7%) | 117 (44.7%) |  |
| Yes | 1,446 (51.4%) | 343 (44.2%) | 550 (51.7%) | 408 (57.3%) | 145 (55.3%) |  |
| BMI category |  |  |  |  |  | <0.001 |
| Underweight | 71 (2.5%) | 34 (4.4%) | 24 (2.3%) | 10 (1.4%) | 3 (1.1%) |  |
| Healthy weight | 1,295 (46.0%) | 354 (45.6%) | 452 (42.5%) | 371 (52.1%) | 118 (45.0%) |  |
| Overweight | 851 (30.3%) | 221 (28.5%) | 322 (30.3%) | 218 (30.6%) | 90 (34.4%) |  |
| Obesity | 596 (21.2%) | 167 (21.5%) | 265 (24.9%) | 113 (15.9%) | 51 (19.5%) |  |
| Daily smoker |  |  |  |  |  | <0.001 |
| No | 2,504 (89.0%) | 666 (85.8%) | 972 (91.4%) | 658 (92.4%) | 208 (79.4%) |  |
| Yes | 309 (11.0%) | 110 (14.2%) | 91 (8.6%) | 54 (7.6%) | 54 (20.6%) |  |
| General health status |  |  |  |  |  | <0.001 |
| Fair | 518 (18.4%) | 192 (24.7%) | 175 (16.5%) | 95 (13.3%) | 56 (21.4%) |  |
| Very good | 1,639 (58.3%) | 424 (54.6%) | 648 (61.0%) | 418 (58.7%) | 149 (56.9%) |  |
| Excellent | 656 (23.3%) | 160 (20.6%) | 240 (22.6%) | 199 (27.9%) | 57 (21.8%) |  |
| Vaccinated for COVID-19 |  |  |  |  |  | <0.001 |
| No | 734 (26.1%) | 312 (40.2%) | 223 (21.0%) | 129 (18.1%) | 70 (26.7%) |  |
| Yes | 2,079 (73.9%) | 464 (59.8%) | 840 (79.0%) | 583 (81.9%) | 192 (73.3%) |  |
| Received mental health diagnosis |  |  |  |  |  | <0.001 |
| No | 487 (17.3%) | 84 (10.8%) | 196 (18.4%) | 159 (22.3%) | 48 (18.3%) |  |
| Yes | 2,326 (82.7%) | 692 (89.2%) | 867 (81.6%) | 553 (77.7%) | 214 (81.7%) |  |
| Received prescription psychiatric treatment in past 4 weeks |  |  |  |  |  | 0.723 |
| No | 2,208 (78.5%) | 607 (78.2%) | 838 (78.8%) | 564 (79.2%) | 199 (76.0%) |  |
| Yes | 605 (21.5%) | 169 (21.8%) | 225 (21.2%) | 148 (20.8%) | 63 (24.0%) |  |
| Received therapy in past 4 weeks |  |  |  |  |  | 0.242 |
| No | 2,271 (80.7%) | 628 (80.9%) | 872 (82.0%) | 557 (78.2%) | 214 (81.7%) |  |
| Yes | 542 (19.3%) | 148 (19.1%) | 191 (18.0%) | 155 (21.8%) | 48 (18.3%) |  |
| Used any drugs in the past month |  |  |  |  |  | <0.001 |
| No | 2,034 (72.3%) | 649 (83.6%) | 805 (75.7%) | 452 (63.5%) | 128 (48.9%) |  |
| Yes | 779 (27.7%) | 127 (16.4%) | 258 (24.3%) | 260 (36.5%) | 134 (51.1%) |  |
| Identifies as being in recovery for drugs and/or alcohol |  |  |  |  |  | <0.001 |
| No | 2,599 (92.4%) | 666 (85.8%) | 1,022 (96.1%) | 681 (95.6%) | 230 (87.8%) |  |
| Yes | 214 (7.6%) | 110 (14.2%) | 41 (3.9%) | 31 (4.4%) | 32 (12.2%) |  |
| Relationship status |  |  |  |  |  | <0.001 |
| Not in a relationship | 997 (35.4%) | 332 (42.8%) | 365 (34.3%) | 199 (27.9%) | 101 (38.5%) |  |
| In a relationship | 1,816 (64.6%) | 444 (57.2%) | 698 (65.7%) | 513 (72.1%) | 161 (61.5%) |  |
| Life satisfaction |  |  |  |  |  | <0.001 |
| Low | 329 (11.7%) | 102 (13.1%) | 115 (10.8%) | 68 (9.6%) | 44 (16.8%) |  |
| Medium | 921 (32.7%) | 240 (30.9%) | 330 (31.0%) | 249 (35.0%) | 102 (38.9%) |  |
| High | 1,563 (55.6%) | 434 (55.9%) | 618 (58.1%) | 395 (55.5%) | 116 (44.3%) |  |
| Social engagement level |  |  |  |  |  | <0.001 |
| Low | 623 (22.1%) | 282 (36.3%) | 202 (19.0%) | 107 (15.0%) | 32 (12.2%) |  |
| Medium | 1,367 (48.6%) | 363 (46.8%) | 542 (51.0%) | 334 (46.9%) | 128 (48.9%) |  |
| High | 823 (29.3%) | 131 (16.9%) | 319 (30.0%) | 271 (38.1%) | 102 (38.9%) |  |
| Strong sense of neighborhood community |  |  |  |  |  | 0.204 |
| Strongly agree | 296 (10.5%) | 93 (12.0%) | 109 (10.3%) | 68 (9.6%) | 26 (9.9%) |  |
| Agree | 837 (29.8%) | 207 (26.7%) | 318 (29.9%) | 228 (32.0%) | 84 (32.1%) |  |
| Neutral | 1,108 (39.4%) | 315 (40.6%) | 424 (39.9%) | 275 (38.6%) | 94 (35.9%) |  |
| Disagree | 404 (14.4%) | 106 (13.7%) | 153 (14.4%) | 109 (15.3%) | 36 (13.7%) |  |
| Strongly disagree | 168 (6.0%) | 55 (7.1%) | 59 (5.6%) | 32 (4.5%) | 22 (8.4%) |  |
| Faith, religion or spirituality gives sense of purpose |  |  |  |  |  | <0.001 |
| Strongly agree | 450 (24.4%) | 194 (34.0%) | 146 (20.9%) | 71 (17.1%) | 39 (24.1%) |  |
| Agree | 610 (33.0%) | 168 (29.5%) | 254 (36.4%) | 148 (35.6%) | 40 (24.7%) |  |
| Neutral | 482 (26.1%) | 138 (24.2%) | 185 (26.5%) | 113 (27.2%) | 46 (28.4%) |  |
| Disagree | 153 (8.3%) | 29 (5.1%) | 65 (9.3%) | 46 (11.1%) | 13 (8.0%) |  |
| Strongly disagree | 151 (8.2%) | 41 (7.2%) | 48 (6.9%) | 38 (9.1%) | 24 (14.8%) |  |

<sup>1</sup>Mean (SD); n (%)
<sup>2</sup>Kruskal-Wallis rank sum test; Pearson's Chi-squared test

**Table 3.** Covariate formats and corresponding model types used in the outcome, covariate, and treatment models^1^.

| Variable name | Independent variable | Type<br>Dependent variable and corresponding model |
| --- | --- | --- |
| Time-fixed covariates |  |  |
| Age | Natural cubic spline function (df=3) | Not predicted |
| Sex | Binary | Not predicted |
| Race/ethnicity | 5 categories | Not predicted |
| Education | 4 categories | Not predicted |
| Body mass index | 4 categories | Not predicted |
| Annual household income | 5 categories | Not predicted |
| Household size | 5 categories | Not predicted |
| Residential area type | 3 categories | Not predicted |
| Daily smoker | Binary | Not predicted |
| Relationship status | Binary | Not predicted |
| Sense of neighborhood community | 5 categories | Not predicted |
| Life satisfaction level | Continuous | Not predicted |
| Religiosity | 5 categories | Not predicted |
| Social engagement activities | Continuous | Not predicted |
| In recovery for drugs or alcohol | Binary | Not predicted |
| Pre-baseline summary: Mean of AUDIT-C score | Continuous | Not predicted |
| Pre-baseline summary: Any high-risk drinking | Binary | Not predicted |
| Pre-baseline summary: Mean GAD-7 score | Continuous | Not predicted |
| Pre-baseline summary: Mean PHQ-8 score | Continuous | Not predicted |
| Pre-baseline summary: Mean physical activity | Continuous | Not predicted |
| Pre-baseline summary: Mean housing insecurity score | Continuous | Not predicted |
| Pre-baseline summary: Any food insecurity | Binary | Not predicted |
| Time-varying covariates |  |  |
| Time | 5-category indicator (included in all time-varying models) | Not predicted |
| Health insurance status | Binary | Logistic regression |
| Housing instability | Binary | Logistic regression |
| Food insecurity | Binary | Logistic regression |
| Receives government food assistance | Binary | Logistic regression |
| General health status | 3 categories | Multinomial logistic regression |
| Chronic conditions diagnosed during follow-up | Binary | Logistic regression |
| Anxiety or depression diagnosis | Binary | Logistic regression |
| Received therapy in past 4 weeks | Binary | Logistic regression |
| Received psychiatric medication in past 4 weeks | Binary | Logistic regression |
| Fully vaccinated for COVID-19 | Binary | Logistic regression |
| Drug use | Binary | Logistic regression |
| Minutes per week of physical activity | Natural cubic spline function (df=3) | Two-part zero-inflated normal |
| Employment status | 3 categories | Multinomial logistic regression |
| Number of poor physical health days (1-30) | Natural cubic spline function (df=3) | Two-part zero-inflated normal |
| Number of poor mental health days (1-30) | Natural cubic spline function (df=3) | Two-part zero-inflated normal |
| Disability status | Binary | Logistic regression |
| PHQ-8 score | Natural cubic spline function (df=3) | Two-part zero-inflated normal |
| GAD-7 score | Natural cubic spline function (df=3) | Two-part zero-inflated normal |
| Moderate/severe depression symptoms (PHQ-8 ≥ 10) | Binary | Logistic regression |
| Moderate/severe anxiety symptoms (GAD-7 ≥ 10) | Binary | Logistic regression |
| Treatment |  |  |
| Alcohol use category | 4 categories | Multinomial logistic regression |
| Outcomes |  |  |
| PHQ-8 score in December 2023 | Continuous | Two-part models (logistic model for zero vs. positive; linear model for positive values; variance modeled as a smooth function of the predicted mean to allow for heteroskedasticity; truncated normal draws for simulation) |
| GAD-7 score in December 2023 |  |  |

<sup>1</sup>*Time-varying covariates and treatment models were specified conditional on all prior covariate and exposure history (lagged values), along with baseline covariates and time; select simplifications were applied where needed to address sparse-data limitations.*

Under the natural course of alcohol use, the estimated prevalence of moderate-to-severe depression symptoms (PHQ-8 ≥10) at the end of follow-up (December 2023) was 12.67% (95% CI: 12.17, 13.23). Under the hypothetical intervention in which moderate to high-risk alcohol consumption was reduced to low-risk levels at each follow-up assessment through September 2023, the estimated prevalence was 12.77% (95% CI: 12.26, 13.29), corresponding to a risk difference of 0.10 percentage points (95% CI: -0.22, 0.50) (Table 4). Similarly, for moderate-to-severe anxiety symptoms (GAD-7 ≥10), the estimated prevalence under the natural course was 10.40% (95% CI: 8.89, 11.89), compared to 10.25% (95% CI: 8.76, 11.98) under the intervention, yielding a risk difference of -0.15 percentage points (95% CI: -0.68, 0.35) (Table 4).

**Table 4.**
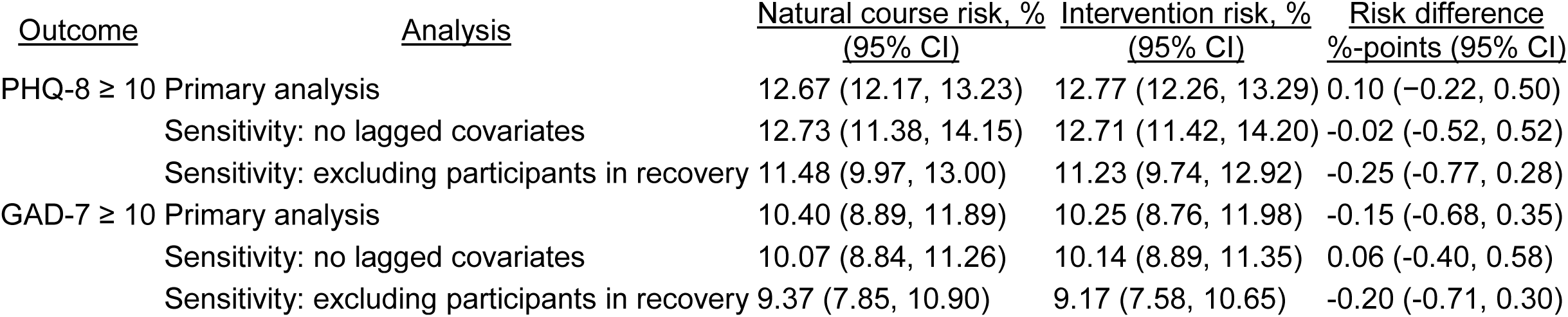
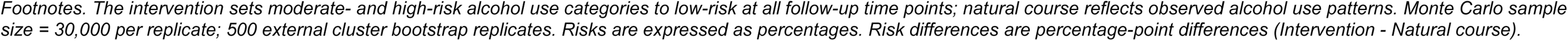
G-computation estimates of risk and risk differences under natural course and sustained low-risk alcohol intervention.

| Outcome | Analysis | Natural course risk, % | Intervention risk, % | Risk difference |
| --- | --- | --- | --- | --- |
|  |  | (95% CI) | (95% CI) | %-points (95% CI) |
| PHQ-8 ≥ 10 | Primary analysis | 12.67 (12.17, 13.23) | 12.77 (12.26, 13.29) | 0.10 (−0.22, 0.50) |
|  | Sensitivity: no lagged covariates | 12.73 (11.38, 14.15) | 12.71 (11.42, 14.20) | −0.02 (−0.52, 0.52) |
|  | Sensitivity: excluding participants in recovery | 11.48 (9.97, 13.00) | 11.23 (9.74, 12.92) | −0.25 (−0.77, 0.28) |
| GAD-7 ≥ 10 | Primary analysis | 10.40 (8.89, 11.89) | 10.25 (8.76, 11.98) | −0.15 (−0.68, 0.35) |
|  | Sensitivity: no lagged covariates | 10.07 (8.84, 11.26) | 10.14 (8.89, 11.35) | 0.06 (−0.40, 0.58) |
|  | Sensitivity: excluding participants in recovery | 9.37 (7.85, 10.90) | 9.17 (7.58, 10.65) | −0.20 (−0.71, 0.30) |

*Footnotes. The intervention sets moderate- and high-risk alcohol use categories to low-risk at all follow-up time points; natural course reflects observed alcohol use patterns. Monte Carlo sample size = 30,000 per replicate; 500 external cluster bootstrap replicates. Risks are expressed as percentages. Risk differences are percentage-point differences (Intervention - Natural course).*

Results were consistent in sensitivity analyses using time-varying covariates measured at the same assessment as alcohol exposure rather than lagged values. Estimated risk differences remained near null for both depression symptoms (-0.02 percentage points; 95% CI: -0.52, 0.52) and anxiety symptoms (0.06 percentage points; 95% CI: -0.40, 0.58) (Table 4), indicating that findings were not sensitive to assumptions about covariate timing. Similarly, after omitting participants in recovery for alcohol or drugs at baseline (N=214), estimated risk differences remained near null for both depression symptoms (-0.25 percentage points; 95% CI: -0.77, 0.28) and anxiety symptoms (-0.20 percentage points; 95% CI: -0.71, 0.30), indicating no evidence that findings were biased by recovery-related abstinence or alcohol reduction at baseline.

The estimated prevalence of moderate-to-severe depression symptoms under the natural course from the parametric g-formula (12.7%) was similar in magnitude to the inverse probability weighted estimate of the natural course under no censoring (14.2%), with an absolute difference of 1.5 percentage points. Additionally, simulated trajectories closely tracked inverse probability weighted observed estimates across timepoints for all modeled covariates, indicating good agreement between the model-based estimates and the observed data (Supplemental Figure S1). These comparisons are commonly used as informal checks of model specification in parametric g-formula analyses.^43^

## Discussion

In this longitudinal cohort of U.S. adults, we used a target trial emulation framework with the parametric g-formula to estimate the impact of a hypothetical intervention reducing alcohol consumption to low-risk levels on the population prevalence of moderate-to-severe anxiety and depression symptoms among individuals without underlying health conditions. Estimated differences between the intervention and natural course were near null for both outcomes and consistent across sensitivity analyses. Notably, even at the extremes of the confidence intervals, differences remained small and did not indicate a meaningful population-level effect. Findings suggest that, at the population level, reducing alcohol consumption alone may be insufficient to meaningfully shift the overall burden of anxiety and depression. Thus, while reducing heavy alcohol consumption is well-established to be important for overall health,^44–46^ population-level interventions focused solely on alcohol consumption may have limited impact on the burden of anxiety and depression symptoms in the population.

In previous analyses of this cohort using causal inference methods to account for time-varying confounding, high-risk alcohol consumption was associated with higher mean anxiety and depression scores at follow-up, whereas moderate-risk drinking did not differ meaningfully from abstinence or low-risk drinking among the same individuals without underlying health conditions.^32^ These findings, consistent with a broader literature linking heavy or hazardous drinking to worse mental health outcomes, suggest that reductions in alcohol use among individuals with high-risk drinking may plausibly improve mental health outcomes within this subgroup.^21,47,48^ However, such subgroup-level effects do not necessarily translate into substantial changes in population-level prevalence of mental health symptoms. One explanation for the limited population-level impact observed in the present study is the distribution of alcohol consumption, whereby individuals with the highest levels of drinking, among whom alcohol use is most strongly associated with anxiety and depression symptoms, represent a relatively small proportion of the population. In this cohort, 9.3% of participants were classified as high-risk at baseline, a proportion that is broadly consistent with the relatively small share of adults engaging in high-risk or heavy drinking in national estimates.^49^ As a result, even if reductions in alcohol use meaningfully improve mental health symptoms among high-risk drinkers, the overall impact on population prevalence of mental health symptoms may be modest.

Attempts to estimate intervention effects within the subgroup of high-risk drinkers were limited by small sample size and resulting model instability, precluding reliable inference in this group. Nevertheless, interventions targeting high-risk drinking remain critical for reducing individual-level mental health symptoms and other well-established alcohol-related harms, including injury, liver disease, cardiovascular disease, and several cancers.^44–46^

Our findings may also be viewed in the context of alcohol’s role as a coping behavior. Alcohol is often used in response to psychological distress and may provide short-term relief of symptoms; however, over time has been shown to exacerbate both mental health symptoms and maladaptive drinking.^50,51^ The observed null findings may reflect that underlying factors that may prompt individuals to cope, such as psychosocial stressors, work-life imbalance, and financial strain, are themselves important drivers of anxiety and depression, and are unlikely to be addressed by reductions in alcohol use alone.^52–54^ Accordingly, reducing alcohol consumption without addressing these underlying drivers, or without the simultaneous replacement of maladaptive coping with more effective strategies, such as physical activity, mindfulness, and mental health treatment, may not lead to measurable improvements in symptoms.^55^

This analysis was designed to estimate the impact of reducing alcohol consumption itself, rather than the full effect of a specific program or policy. Real-world interventions may also affect mental health through other pathways. For example, alcohol use is often closely tied to social activity, and an intervention that reduces drinking by limiting participation in alcohol-centered settings, without replacing those opportunities for connection, could have unintended negative effects on mental health.^56,57^ In contrast, interventions that reduce alcohol use while maintaining or strengthening social connection could have more favorable effects. Therefore, our near-null estimate should not be interpreted to mean that all alcohol-reduction programs would have no impact on mental health; their effects may differ depending on whether they also disrupt or strengthen social connection and other sources of support.

This analysis extends prior work by estimating the population-level impact of a defined intervention on alcohol consumption, rather than describing differences across drinking groups. By applying the parametric g-formula within a target trial emulation framework, we were able to estimate mental health symptoms under sustained reductions to low-risk drinking, addressing sources of bias that commonly affect conventional analyses where alcohol is the exposure, including health-related selection into drinking, reverse causation, and time-varying confounding.^24,25,58^ This approach allows evaluation of questions that are directly relevant to public health decision-making; namely, how population mental health might change under realistic reductions in alcohol consumption.^31^ In contrast, conventional analyses comparing alcohol consumption groups, often using abstainers as a reference, describe differences between groups but do not estimate the effects of changing drinking behavior. By focusing on reductions among individuals exceeding low-risk levels, this analysis provides a more policy-relevant assessment of intervention effects.

The intervention evaluated in this study was defined using AUDIT-C categories, which capture clinically meaningful patterns of alcohol use based on frequency, quantity, and binge drinking and align with screening and brief intervention frameworks commonly used in clinical and public health settings.^59–61^ However, because AUDIT-C categories summarize multiple dimensions of drinking behavior, “low-risk” does not correspond to a single, specific consumption threshold. Accordingly, our estimates should be interpreted as reflecting shifts toward lower-risk drinking patterns rather than adherence to a specific behavioral recommendation. Future studies may clarify the impact of adherence to guideline-defined drinking thresholds based on daily or weekly consumption.

### Strengths and limitations

This study offers several key strengths. First, the CHASING COVID Cohort provides a large, diverse U.S. sample with longitudinal data spanning three years. Second, the application of g-computation represents a methodological advancement to current literature, allowing for the estimation of outcomes under sustained alcohol-use strategies while adjusting for time-varying confounding and loss to follow-up. Finally, by incorporating time-varying covariates related to material hardship, physical activity, drug/alcohol recovery, and social connectedness, we enhanced confounding control beyond previous studies.

There are various limitations to note. First, the analytic sample excluded individuals with underlying documented physical health conditions at baseline, and findings may not generalize to populations with poorer baseline health, and results may differ in these populations. Second, alcohol consumption and mental health symptoms were self-reported and may be subject to measurement error, which could attenuate associations if alcohol use or symptom severity were underreported. In addition, the intervention evaluated in this study modeled reductions in alcohol use in isolation; in practice, changes in drinking behavior may occur alongside other behavioral or clinical interventions, and their combined effects remain unexamined. Our analysis also estimated population-level effects under sustained intervention strategies and did not directly examine within-individual changes in alcohol use and mental health symptoms over time; future work using within-person approaches may help clarify the temporal relationship between changes in drinking behavior and changes in symptom burden. Third, although the parametric g-formula appropriately adjusts for measured time-varying confounding, its validity depends on correct specification of models and the absence of unmeasured confounding.^30^ As with all observational data, relevant confounders may not be fully captured by available covariates. For example, personality characteristics such as extraversion, conscientiousness, and agreeableness, which may influence both alcohol use and mental health, were not measured and could contribute to residual confounding.^62^

### Conclusion

Findings from this study suggest that population-level reductions in alcohol consumption to low-risk levels may have limited impact on the population-prevalence of anxiety and depression symptoms, with no evidence of either decreased or increased symptom burden. While alcohol use is an important modifiable risk factor for a range of adverse health outcomes, reducing alcohol use alone may not meaningfully shift the population burden of common mental health disorders. Efforts to improve population mental health may therefore require addressing a broader set of behavioral, social, and structural determinants driving common mental health symptoms in addition to alcohol use.

## Data Availability

The datasets generated and/or analyzed during the current study are available in the repository, Zenodo: DOI: 10.5281/zenodo.6127734. Some data elements are not publicly available due to funder requirements, but are available from the authors upon reasonable request, subject to approval and available resources.

https://zenodo.org/records/7305435

## Acknowledgements

We thank the participants of the Communities, Households, and SARS-CoV-2 Epidemiology COVID Cohort Study for their contribution to the advancement of science.

## Funding Sources

This work was supported by the National Institute of Allergy and Infectious Diseases (NIAID), award number UH3AI133675 (MPIs: D Nash and C Grov), National Institute of Mental Health (NIMH) award RF1MH132360 (MPIs: D Nash and A Parcesepe), National Institute of Child Health and Human Development grant P2C HD050924 (Carolina Population Center), Pfizer Inc., the CUNY Institute for Implementation Science in Population Health (cunyisph.org), the CUNY Graduate School of Public Health and Health Policy Department of Epidemiology and Biostatistics, and the COVID-19 Grant Program of the CUNY Graduate School of Public Health and Health Policy. The funders played no role in the production of this manuscript or necessarily endorse the findings.

## CRediT Author Statement

**Jenna Sanborn:** Conceptualization, Methodology, Software, Formal analysis, Investigation, Writing-Original Draft. **Denis Nash**: Writing-Review & Editing, Funding acquisition. **McKaylee Robertson:** Writing-Review & Editing. **Angela M Parcesepe**: Writing-Review & Editing, Funding acquisition. **Zachary Shahn:** Methodology, Writing-Review and Editing, Supervision.

## Notes

### Competing Interest Statement

DN received consulting fees from Abbvie and Gilead and has a research grant from Pfizer for his institution (CUNY SPH). The other authors declare no conflict of interest. The funders had no role in the design of this study, in the collection, analysis, or interpretation of data, in the writing of this manuscript, or in the decision to publish these results.

### Author Declarations

The Institutional Review Board at CUNY Graduate School of Public Health gave ethical approval of this work

