## Supplementary Material for "Prevalence of anxiety and depression symptoms under population-level alcohol reduction to low-risk drinking: A parametric g-computation analysis in a U.S. nationwide cohort"

| **Supplementary Table S1. Comparison of baseline participant characteristics between the analytic sample, those with underlying conditions, and the full CHASING COVID Cohort, September 2021** | | | |
| --- | --- | --- | --- |
|  | **Full Cohort** | **Without underlying conditions (analytic sample)** | **With underlying conditions or poor self-rated health** |
| **Characteristic** | **N = 4,673**^1^ | **N = 2,794**^1^ | **N = 1,860**^1^ |
| **Moderate/severe depression Symptoms** |  |  |  |
| Yes | 1,097 (23.5%) | 566 (20.3%) | 527 (28.3%) |
| No | 3,576 (76.5%) | 2,228 (79.7%) | 1,333 (71.7%) |
| **Moderate/severe anxiety symptoms** |  |  |  |
| Yes | 902 (19.3%) | 494 (17.7%) | 406 (21.8%) |
| No | 3,771 (80.7%) | 2,300 (82.3%) | 1,454 (78.2%) |
| **Alcohol consumption level** |  |  |  |
| Abstinent | 1,418 (30.3%) | 769 (27.5%) | 642 (34.5%) |
| Low risk | 1,748 (37.4%) | 1,056 (37.8%) | 685 (36.8%) |
| Moderate risk | 1,069 (22.9%) | 709 (25.4%) | 357 (19.2%) |
| High/severe risk | 438 (9.4%) | 260 (9.3%) | 176 (9.5%) |
| **Age (years)** | 42.72 (15.09) | 39.50 (13.91) | 47.59 (15.51) |
| **Sex** |  |  |  |
| Male | 2,117 (45.3%) | 1,193 (42.7%) | 918 (49.4%) |
| Female | 2,556 (54.7%) | 1,601 (57.3%) | 942 (50.6%) |
| **Race/Ethnicity** |  |  |  |
| Hispanic | 749 (16.0%) | 472 (16.9%) | 272 (14.6%) |
| White (non-Hispanic) | 2,973 (63.6%) | 1,741 (62.3%) | 1,224 (65.8%) |
| Black (non-Hispanic) | 447 (9.6%) | 229 (8.2%) | 214 (11.5%) |
| Asian/Pacific Islander (non-Hispanic) | 342 (7.3%) | 268 (9.6%) | 73 (3.9%) |
| Other (non-Hispanic) | 162 (3.5%) | 84 (3.0%) | 77 (4.1%) |
| **Education** |  |  |  |
| Less than high school | 68 (1.5%) | 32 (1.1%) | 35 (1.9%) |
| High school | 446 (9.5%) | 232 (8.3%) | 211 (11.3%) |
| Some college | 1,180 (25.3%) | 640 (22.9%) | 534 (28.7%) |
| College graduate | 2,979 (63.7%) | 1,890 (67.6%) | 1,080 (58.1%) |
| **Household size** |  |  |  |
| 1 | 1,180 (25.3%) | 653 (23.4%) | 523 (28.1%) |
| 2 | 1,528 (32.7%) | 891 (31.9%) | 633 (34.0%) |
| 3 | 731 (15.6%) | 445 (15.9%) | 282 (15.2%) |
| 4 | 661 (14.1%) | 436 (15.6%) | 223 (12.0%) |
| 5+ | 573 (12.3%) | 369 (13.2%) | 199 (10.7%) |
| **Residential area type** |  |  |  |
| Suburban/Town | 1,228 (26.3%) | 727 (26.0%) | 496 (26.7%) |
| Rural | 1,400 (30.0%) | 774 (27.7%) | 621 (33.4%) |
| Urban | 2,045 (43.8%) | 1,293 (46.3%) | 743 (39.9%) |
| **Annual household income** |  |  |  |
| <$35,000 | 1,252 (26.8%) | 662 (23.7%) | 583 (31.3%) |
| $35,000–$49,999 | 537 (11.5%) | 300 (10.7%) | 233 (12.5%) |
| $50,000–$69,999 | 713 (15.3%) | 419 (15.0%) | 291 (15.6%) |
| $70,000–$99,000 | 804 (17.2%) | 496 (17.8%) | 308 (16.6%) |
| $100,000+ | 1,367 (29.3%) | 917 (32.8%) | 445 (23.9%) |
| **Employment status** |  |  |  |
| Employed | 3,172 (67.9%) | 2,062 (73.8%) | 1,100 (59.1%) |
| Unemployed | 383 (8.2%) | 176 (6.3%) | 204 (11.0%) |
| Not in labor force | 1,118 (23.9%) | 556 (19.9%) | 556 (29.9%) |
| **Lost income in previous month** |  |  |  |
| No | 4,096 (87.7%) | 2,512 (89.9%) | 1,568 (84.3%) |
| Yes | 577 (12.3%) | 282 (10.1%) | 292 (15.7%) |
| **Has health insurance** |  |  |  |
| No | 464 (9.9%) | 293 (10.5%) | 166 (8.9%) |
| Yes | 4,209 (90.1%) | 2,501 (89.5%) | 1,694 (91.1%) |
| **Housing insecurity** |  |  |  |
| No | 3,399 (72.7%) | 2,122 (75.9%) | 1,270 (68.3%) |
| Yes | 1,274 (27.3%) | 672 (24.1%) | 590 (31.7%) |
| **Food insecurity** |  |  |  |
| No | 3,761 (80.5%) | 2,357 (84.4%) | 1,394 (74.9%) |
| Yes | 912 (19.5%) | 437 (15.6%) | 466 (25.1%) |
| **Received government food assistance** |  |  |  |
| No | 3,514 (75.2%) | 2,209 (79.1%) | 1,297 (69.7%) |
| Yes | 1,159 (24.8%) | 585 (20.9%) | 563 (30.3%) |
| **Physically active (60+ mins per week)** |  |  |  |
| No | 1,828 (39.1%) | 968 (34.6%) | 849 (45.6%) |
| Yes | 2,845 (60.9%) | 1,826 (65.4%) | 1,011 (54.4%) |
| **BMI category** |  |  |  |
| Underweight | 99 (2.1%) | 72 (2.6%) | 27 (1.5%) |
| Healthy weight | 1,744 (37.3%) | 1,285 (46.0%) | 453 (24.4%) |
| Overweight | 1,411 (30.2%) | 850 (30.4%) | 554 (29.8%) |
| Obesity | 1,419 (30.4%) | 587 (21.0%) | 826 (44.4%) |
| **Has chronic health condition** |  |  |  |
| No | 2,881 (61.7%) | 2,794 (100.0%) | 68 (3.7%) |
| Yes | 1,792 (38.3%) | 0 (0.0%) | 1,792 (96.3%) |
| **Daily smoker** |  |  |  |
| No | 4,041 (86.5%) | 2,493 (89.2%) | 1,536 (82.6%) |
| Yes | 632 (13.5%) | 301 (10.8%) | 324 (17.4%) |
| **General health status** |  |  |  |
| Excellent | 107 (2.3%) | 7 (0.3%) | 100 (5.4%) |
| Good | 1,178 (25.2%) | 492 (17.6%) | 681 (36.6%) |
| Fair | 2,489 (53.3%) | 1,596 (57.1%) | 882 (47.4%) |
| Poor | 899 (19.2%) | 699 (25.0%) | 197 (10.6%) |
| **Vaccinated for COVID-19** |  |  |  |
| No | 660 (14.1%) | 417 (14.9%) | 242 (13.0%) |
| Yes | 4,013 (85.9%) | 2,377 (85.1%) | 1,618 (87.0%) |
| **Received mental health diagnosis** |  |  |  |
| No | 789 (16.9%) | 484 (17.3%) | 303 (16.3%) |
| Yes | 3,884 (83.1%) | 2,310 (82.7%) | 1,557 (83.7%) |
| **Received mental health treatment** |  |  |  |
| No | 3,081 (81.6%) | 1,946 (85.8%) | 1,124 (75.3%) |
| Yes | 693 (18.4%) | 321 (14.2%) | 369 (24.7%) |
| **Used any drugs in the past month** |  |  |  |
| No | 3,358 (71.9%) | 2,022 (72.4%) | 1,324 (71.2%) |
| Yes | 1,315 (28.1%) | 772 (27.6%) | 536 (28.8%) |
| **Identifies as being in recovery for drugs and/or alcohol** |  |  |  |
| No | 4,054 (86.8%) | 2,483 (88.9%) | 1,556 (83.7%) |
| Yes | 619 (13.2%) | 311 (11.1%) | 304 (16.3%) |
| **Relationship status** |  |  |  |
| Not in a relationship | 1,752 (37.5%) | 966 (34.6%) | 779 (41.9%) |
| In a relationship | 2,921 (62.5%) | 1,828 (65.4%) | 1,081 (58.1%) |
| **Life satisfaction** |  |  |  |
| Low | 626 (13.4%) | 333 (11.9%) | 290 (15.6%) |
| Medium | 1,552 (33.2%) | 920 (32.9%) | 624 (33.5%) |
| High | 2,495 (53.4%) | 1,541 (55.2%) | 946 (50.9%) |
| **Social engagement level** |  |  |  |
| Low | 1,066 (22.8%) | 555 (19.9%) | 502 (27.0%) |
| Medium | 2,286 (48.9%) | 1,376 (49.2%) | 903 (48.5%) |
| High | 1,321 (28.3%) | 863 (30.9%) | 455 (24.5%) |
| **Strong sense of neighborhood community** |  |  |  |
| Strongly agree | 517 (11.1%) | 293 (10.5%) | 220 (11.8%) |
| Agree | 1,408 (30.1%) | 832 (29.8%) | 573 (30.8%) |
| Neutral | 1,780 (38.1%) | 1,104 (39.5%) | 670 (36.0%) |
| Disagree | 674 (14.4%) | 397 (14.2%) | 274 (14.7%) |
| Strongly disagree | 294 (6.3%) | 168 (6.0%) | 123 (6.6%) |
| **Faith, religion or spirituality gives sense of purpose** |  |  |  |
| Strongly agree | 801 (25.4%) | 446 (24.3%) | 352 (27.0%) |
| Agree | 1,060 (33.6%) | 604 (32.9%) | 452 (34.6%) |
| Neutral | 815 (25.8%) | 479 (26.1%) | 330 (25.3%) |
| Disagree | 240 (7.6%) | 155 (8.4%) | 85 (6.5%) |
| Strongly disagree | 239 (7.6%) | 152 (8.3%) | 86 (6.6%) |
| ^1^Mean (SD); n (%) | | | |

**Supplemental Methods: Statistical Analysis**

### 1.1. ****Parametric g-formula implementation****

We implemented the parametric g-formula using Monte Carlo simulation to estimate the effect of a sustained alcohol reduction intervention on end-of-follow-up mental health outcomes. The intervention corresponded to a threshold intervention in which alcohol consumption was set to low-risk levels at each follow-up assessment when the observed value exceeded this threshold, and otherwise allowed to follow its natural course absent any intervention. This class of interventions whereby the intervention corresponds to a threshold intervention that depends on the natural value of treatment, can be estimated using the extended g-formula, which requires specification of models for the conditional distribution of treatment, time-varying covariates, and outcomes.^1^ Simulations were conducted using 30,000 individuals to approximate the distribution of outcomes under both the natural course and intervention scenarios.

### 1.2. ****Model specification****

Regression models were specified for alcohol use, time-varying covariates, and outcomes conditional on prior exposure and covariate history, time, and baseline covariates. Model forms were selected based on the distribution of each variable (e.g., categorical, normal, truncated normal). Continuous predictors were modeled using natural cubic splines to allow for nonlinear relationships.

The treatment model for alcohol use included prior alcohol use, time-varying confounders, and baseline covariates. Time-varying covariate models included prior exposure and covariate history as predictors. Selected simplifications were applied where necessary to address sparse data limitations.

### 1.3. ****Censoring and model diagnostics****

Participants were censored at the first missing alcohol assessment after baseline, such that person-time was contributed only up to the last observed alcohol measurement. The g-formula accounts for informative censoring by estimating outcomes under a scenario in which all participants remain under observation throughout follow-up.

To assess model fit, we compared inverse probability of censoring weighted estimates of observed covariate distributions over follow-up with corresponding distributions simulated under the natural course. The censoring process was modeled using pooled logistic regression for the probability of remaining uncensored at each time point, conditional on time, prior mental health symptoms (PHQ-8 and GAD-7 scores), time-varying covariates, and baseline sociodemographic characteristics.

### 1.4. ****Simulation of end-of-follow-up outcomes****

End-of-follow-up depression and anxiety symptom scores were simulated under both the natural course and intervention scenarios. Because PHQ-8 and GAD-7 scores are semi-continuous with a substantial mass at zero, we used a two-part modeling approach, with a logistic model for the probability of a zero score and a continuous model for positive values. Simulated scores were then used to estimate the risk of moderate-to-severe symptoms, defined as PHQ-8 ≥10 and GAD-7 ≥10.

### 1.5. ****Bootstrapping****

Ninety-five percent confidence intervals were estimated using a nonparametric cluster bootstrap with resampling at the participant level. For each of 500 bootstrap replicates, participants were sampled with replacement, models were refit, and the g-formula was approximated via Monte Carlo simulation. Percentile-based confidence intervals were calculated from the bootstrap distribution of the natural course risk, intervention risk, and risk difference.

### 1.6. ****Sensitivity analyses****

Two sensitivity analyses were conducted for each outcome. First, to assess the robustness of our findings to covariate timing, analyses were repeated using time-varying covariates measured at the same assessment as alcohol exposure rather than lagged values from the prior assessment. While the primary analysis used lagged covariates to ensure temporal ordering, this approach allowed covariates to align more closely with exposure measurement and may better control for time-varying confounding, albeit under the assumption that covariates preceded exposure within the same assessment.

Second, to assess potential bias related to recovery-related changes in alcohol use, we repeated the analyses after excluding participants who reported being in recovery from alcohol or drug use at analytic baseline, as these individuals may have reduced or stopped drinking prior to study enrollment and their prior drinking history would not be captured in the cohort data.

**Supplementary Figure S1. Comparison of inverse probability weighted observed and g-formula-simulated covariate trajectories under the natural course**

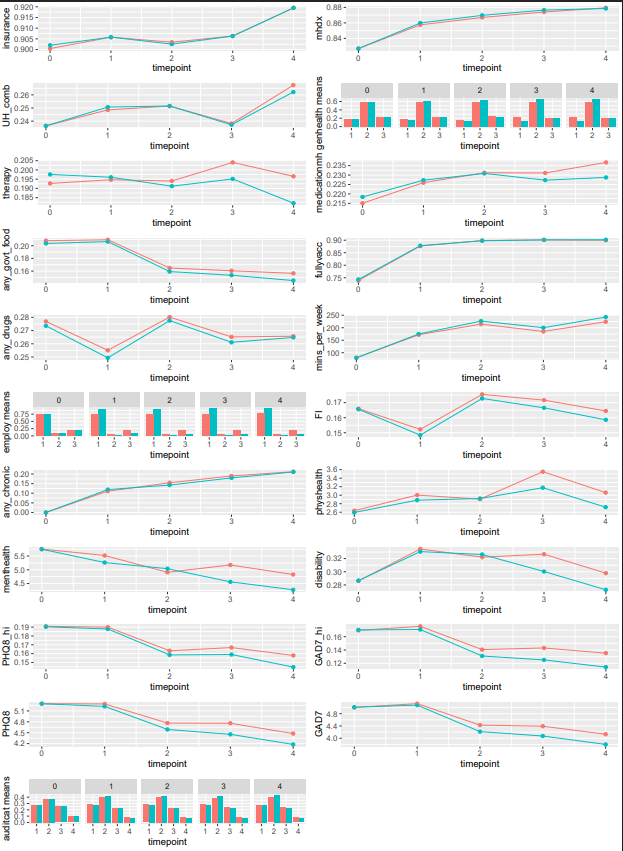

| **Supplementary Table S2. Definitions and assessment timing of variables used in analysis** | | | | |
| --- | --- | --- | --- | --- |
| Variable | Variable Type | Definition | Assessment Timing | Assessment Question(s) |
| Exposure & Outcomes | | | | |
| Alcohol consumption risk level* | Categorical | Calculated by summing scores from three assessment questions; Each has five answer choices, with point values ranging from 0 to 4. The final score ranges from 0 to 12, and is used to determine alcohol consumption risk level | Sept 2021, Dec 2021,  Mar 2022, Oct 2022, Apr 2023, Sept 2023 | In the past month, how often do you have a drink containing alcohol? [Never, Monthly or less, 2-4 times a month, 2-3 times a week, 4 or more times a week]  *If do not drink alcohol, then skip:* In the last month, how many standard drinks containing alcohol do you have on a typical day? [1 or 2, 3 or 4, 5 or 6, 7-9, 10 or more]  *If do not drink alcohol, then skip:* In the last month, how often do you have six or more drinks on one occasion? [Never, Less than monthly, Monthly, Weekly, Daily or almost daily] |
| GAD-7 score (anxiety)* | Continuous | Sum of scores where not at all =0, several days=1, over half the days=2 and nearly every day=3. The final score ranges from 0-21. | Dec 2023 | In the past month, how often have you been bothered by the following symptoms? [Not at all, Several days, Over half the days, Nearly every day]: 1) Feeling nervous, anxious or on edge, 2) Not being able to stop or control worrying, 3) Worrying too much about different things, 4) Trouble relaxing, 5) Being so restless it’s hard to sit still, 6) Becoming easily annoyed or irritable, 7) Feeling afraid as if something awful might happen |
| PHQ-8 score (depression)* | Continuous | Sum of scores, where not at all =0, several days=1, over half the days=2 and nearly every day=3. The final score ranges from 0-24. | Dec 2023 | In the past month, how often have you been bothered by the following symptoms? [Not at all, Several days, Over half the days, Nearly every day]: 1) Feeling down, depressed or hopeless, 2) Trouble falling or staying asleep, or sleeping too much, 3) Feeling tired or having little energy, 4) Poor appetite or overeating, 5) Feeling bad about yourself- or that you are a failure or have let yourself or your family down, 6) Trouble concentrating on things, such as, reading the newspaper or watching television, 7) Moving or speaking so slowly that other people have noticed? Or the opposite- being so fidgety or restless that you have been moving around a lot more than usual |
| Time-fixed covariates | | | | |
| Pre-baseline AUDIT-C scores | Continuous | Average of pre-baseline AUDIT-C scores | Enrollment,  Nov 2020,  May 2021 | In the past month, how often do you have a drink containing alcohol? [Never, Monthly or less, 2-4 times a month, 2-3 times a week, 4 or more times a week]  *If do not drink alcohol, then skip:* In the last month, how many standard drinks containing alcohol do you have on a typical day? [1 or 2, 3 or 4, 5 or 6, 7-9, 10 or more]  *If do not drink alcohol, then skip:* In the last month, how often do you have six or more drinks on one occasion? [Never, Less than monthly, Monthly, Weekly, Daily or almost daily] |
| Age | Continuous | Self-reported age | Enrollment | What is your age? Age ____ |
| Sex at birth | Binary | Collapsed to 2 categories: ‘Male’ and ‘Female’. Missing values, imputed with gender identity, assessed at baseline, if male or female was endorsed | April 2023 | What sex were you assigned at birth? a) Male, b) Female, c) None of the above |
| Race/ethnicity | Categorical | Self-reported race/ethnicity grouped into five categories: Hispanic, White (non-Hispanic), Black (non-Hispanic), Asian/ Pacific Islander (non-Hispanic), Other | Enrollment | Are you Hispanic, Latino/a, or Spanish origin? [Yes, No, Don’t know / Not sure]  Which of these groups would you say best represents your race? Please select all that apply. [Black or African American, American Indian or Alaska Native, Asian, Pacific Islander, White, Other ___, Don’t know/ Not sure] |
| Education | Categorical | Highest education level completed: [Less than high school, high school graduate, some college, college graduate] | Enrollment | What is the highest grade or year of school you completed? [Less than a high school diploma, Grade 12 or GED (High school graduate), College 1 year to 3 years (Some college or technical school), College 4 years or more (College graduate) |
| Annual Household Income | Categorical | Annual household income from all sources: [<$35,000, $50,000-$69,999, $70,000-99,999, $100,000+] | Enrollment | Is your annual household income from all sources: [<$25,000, $25,000-$34,999, $35,000-$49,999, $50,000-$69,999, $70,000-$99,999, $100,000-$149,000, $150,000+] |
| Body Mass Index | Categorical | Calculated as kg/m^2^ based on height and weight and categorized as Underweight, Healthy weight, Overweight, Obesity [ref] | Enrollment | How much do you weigh without shoes? Please answer in pounds ____  About how tall are you without shoes? Please answer in feet and inches. ____ Feet ____ Inches |
| Cigarette smoking status | Binary | Coded as ‘1’ if every day selected | Enrollment | Do you currently smoke cigarettes every day, some days or not at all? (Cigarettes does not include electronic products such as: e-cigarettes, vape pens, personal vaporizers, e-cigars, epipes, e-hookahs, hookah pens, and mods). a) Every day, b) Some days, c) Not at all, d) Don’t know / Not sure |
| Household size | Categorical | Sum total of number of people in each age group (children, 18-59 and 60+) | Enrollment | How many members of your household, including yourself, are between 18-59 years of age? [___number, no one aged 18-59]  How many members of your household, including yourself, are 60 years old or older? [___number, no one aged 60+]  How many children less than 18 years of age live in your household? [___number, no children < 18 live in my household] |
| Residential area type | Categorical | Residential area designation based on zip code categorized as “Suburban/Town”, ‘Rural” and “Urban” | Enrollment | ZIP code was used to create the geographic-level variable for residential area type, assigned based on the NCES Education Demographic and Geographic Estimates locale definitions, using the ZCTA locale file to map ZIP codes to 'Rural', 'Suburban', 'Urban', and 'Town'. Given the low number of 'Town' designations (n=7), ‘Suburban’ and 'Town’ were collapsed into a single category. |
| Neighborhood community | Categorical | Sense of neighborhood community [Strongly agree/agree, Neutral, Stronger disagree/disagree] | Feb 2021 | Living in my current neighborhood gives me a strong sense of community. [Strongly agree, Agree, Neutral, Disagree, Strongly Disagree] |
| Life satisfaction | Continuous | Level of satisfaction with life as a whole | Feb 2021 | Overall, how satisfied are you with life as a whole these days? (0= not satisfied at all, 10= completely satisfied) |
| Spirituality/ Religion | Categorical | Faith, religion, spirituality gives sense of purpose [Strongly agree/agree, Neutral, Stronger disagree/disagree] | Feb 2021 | My faith, religious, or spiritual beliefs give me a sense of direction and purpose in my life. [Strongly agree, Agree, Neutral, Disagree, Strongly disagree] |
| Relationship status | Binary | Classify responses into two categories: ‘Yes’ vs. ‘No/ Don’t know’ | May 2021 | Are you currently in a relationship or seeing someone? [Yes, No, Don’t know/ Not sure] |
| Social engagement level | Categorical | Calculated number of social activities participants spent time doing during the past month: 1) Gathering of 10+ people, either indoors or outdoors, 2) Spent time inside of a house that is not your own, 3) Spent time inside a restaurant or bar, 4) Spent time in the patio or outdoor space of a restaurant or bar, or 5) Spent time at any of the following: The inside of a restaurant or bar, A patio or outdoor space at a restaurant or bar, An indoor movie theatre, A shopping mall, A church, synagogue, mosque or other place of worship, The inside of a house that is not your own, an overnight stay at the residence of family or friends.  Responses categorized as ‘low’, ‘medium’ and ‘high’: low= <2, medium: 2-4, high: 5+ | May 2021 | In the past month, have you gathered in groups with 10 or more people? a) Yes, indoors only, b) Yes, outdoors only, c) Yes, indoors and outdoors, d) No, e) Don’t know/ Not sure  In the past month, have you done any of the following? [Yes, No, Not applicable]  a) Spent time inside of a house that is not your own, b) Spent time inside a restaurant or bar, c) Spent time in the patio or outdoor space of a restaurant or bar, d) Had an overnight stay at a hotel, short-term rental or residence of family or friends  In the past month, have you spent time in any of the following places? Please select all that apply.  a) A hairdresser, salon or barber, b) The inside of a restaurant or bar, c) A patio or outdoor space at a restaurant or bar, d) An indoor movie theatre, e) A shopping mall, f) A church, synagogue, mosque or other place of worship, g) The inside of a house that is not your own, h) A public swimming area such as a pool, lake, ocean or bay, i) a public park, j) a mass gathering like a demonstration or public protest, k) A mass gathering like a political rally, l) a hotel or other short-term rental (like Airbnb) where people outside of your household are staying, m) An overnight stay at the residence of family or friends, n) An overnight trip to another town or city, o) A gym or exercise facility, p) None of the above |
| Substance use or alcohol recovery | Binary | Responses collapsed to two categories: “Yes- currently or previously” vs. “Never/ Not sure” | Enrollment, May 2021, Sept 2021 | Do you identify as being in recovery from drugs? [Yes, I am currently in recovery from drugs, I am not currently in recovery from drugs, but I have previously been in recovery from drugs, I have never been in recovery from drugs, Don’t know/ Not sure]  Do you identify as being in recovery from alcohol? [Yes, I am currently in recovery from alcohol, I am not currently in recovery from alcohol, but I have previously been in recovery from alcohol, I have never been in recovery from alcohol, Don’t know/ Not sure] |
| Time-Varying Covariates | | | | |
| GAD-7 score  (anxiety) | Continuous | Sum of scores where not at all =0, several days=1, over half the days=2 and nearly every day=3. The final score ranges from 0-21. | Pre-baseline (avg from assessments between enrollment and May 2020), Sept 2021, Dec 2021, Mar 2022, Jun 2022, Oct 2022, Dec 2022, Apr 2023, Jun 2023, Sept 2023 | In the past month, how often have you been bothered by the following symptoms? [Not at all, Several days, Over half the days, Nearly every day]: 1) Feeling nervous, anxious or on edge, 2) Not being able to stop or control worrying, 3) Worrying too much about different things, 4) Trouble relaxing, 5) Being so restless it’s hard to sit still, 6) Becoming easily annoyed or irritable, 7) Feeling afraid as if something awful might happen |
| PHQ-8 score (depression) | Continuous | Sum of scores, where not at all =0, several days=1, over half the days=2 and nearly every day=3. The final score ranges from 0-24. | Pre-baseline (avg from assessments between July 2020 and May 2020), Sept 2021, Dec 2021, Mar 2022, Jun 2022, Oct 2022, Dec 2022, Apr 2023, Jun 2023, Sept 2023 | In the past month, how often have you been bothered by the following symptoms? [Not at all, Several days, Over half the days, Nearly every day]: 1) Feeling down, depressed or hopeless, 2) Trouble falling or staying asleep, or sleeping too much, 3) Feeling tired or having little energy, 4) Poor appetite or overeating, 5) Feeling bad about yourself- or that you are a failure or have let yourself or your family down, 6) Trouble concentrating on things, such as, reading the newspaper or watching television, 7) Moving or speaking so slowly that other people have noticed? Or the opposite- being so fidgety or restless that you have been moving around a lot more than usual |
| Physical activity | Continuous | Minutes of physical activity per week calculated using the three assessment questions | Oct 2020, Feb 2021, May 2021, Sept 2021, Dec 2021, Mar 2022, Jun 2022, Oct 2022, Dec 2022, Apr 2023, Jun 2023, Sept 2023 | 1. During the past month, other than your regular job, did you participate in any physical activities or exercises such as running, calisthenics, golf, gardening or walking for exercise? [Yes, No, Don’t know/ Not sure]  2. How many times per week or per month did you take part in this activity during the past month? [__ times per week, __ times per month, don’t know/ not sure]  3. And when you took part in this activity, for how many minutes or hours did you usually keep at it? [__ Number of hours, __ Number of minutes, Don’t know/ Not sure] |
| Received therapy in the past 4 weeks | Categorical | Received counseling from a mental health professional in the previous 4 weeks [Yes vs. No/don’t know] | May 2021, Sept 2021, Dec 2021, Mar 2022, Oct 2022, Apr 2023, Sept 2023 | In the past four weeks, have you received counseling or therapy from a mental health professional, such as a psychologist, social worker, or other licensed mental health professional? [Yes, No, Don’t know/ Not sure] |
| Received psychiatric medication in the past 4 weeks |  | Took prescription medication for mental health in the previous 4 weeks [Yes vs. No/don’t know] | May 2021, Sept 2021, Dec 2021, Mar 2022, Oct 2022, Apr 2023, Sept 2023 | In the past four weeks, have you taken prescription medication for your mental health? [Yes, No, Don’t know/ Not sure] |
| Anxiety or depression diagnosis | Binary | Diagnosed with anxiety, depression or PTSD (yes vs. no) | Enrollment,  Dec 2021, Sept 2023 | Has a doctor, nurse, or other health professional ever told you that you had any of the following? *Select all that apply* [Anxiety, depression, PTSD] |
| Health insurance status | Binary | Participants categorized as having insurance vs. not (yes vs. no/don’t know). | Enrollment, Feb 2021, May 2021,  Sept 2021, Dec 2021,  Mar 2022, Jun 2022,  Oct 2022, Dec 2022,  Apr 2023, Jun 2023,  Sept 2023 | Do you have any kind of health care coverage, including health insurance, prepaid plans such as HMOs, or government plans such as Medicare, or Indian Health Service? [Yes, No, Don’t know/ Not sure] |
| Housing instability | Binary | Categorize as housing insecure if “always” or “usually” is selected | Enrollment, Feb 2021, May 2021, Sept 2021, Dec 2021, Mar 2022, Oct 2022, Apr 2023, Sept 2023 | How often in the past month would you say you were worried or stressed about having enough money to pay your rent/mortgage? [Always, Usually, Sometimes, Rarely, Never] |
| Food insecurity | Binary | Categorize as food insecure if “Often” or “Sometimes” is selected for either of these statements^2^ | Enrollment, Feb 2021, May 2021,  Sept 2021, Dec 2021,  Mar 2022, Oct 2022, Apr 2023, Sept 2023 | For each of the following, you will answer whether the statement was often true, sometimes true, or never true for (you/your household) in the past month.  1) “We worried whether our food would run out before we got money to buy more.” [Often true, sometimes true, never true]  2) “The food that we bought just didn’t last, and we didn’t have money to get more.” Was that often, sometimes or never true for you in the past month? [Often true, sometimes true, never true] |
| Employment status | Categorical | Responses classified into three categories: [Employed, Unemployed, Not in labor force] | Enrollment, Feb 2021, May 2021, Sept 2021, Dec 2021, Mar 2022, Jun 2022, Oct 2022, Dec 2022, Apr 2023, Jun 2023, Sept 2023 | Are you currently…? [Employed for wages, Self-employed, Out of work for less than 1 year, Out of work for 1 year or more, A homemaker, A student, Retired] |
| General health status | Categorical | General health categories: [Poor, Fair, Very Good, Excellent] | July 2020, Feb 2021, May 2021, Sept 2021, Dec 2021, Mar 2022, Jun 2022, Oct 2022, Dec 2022, Apr 2023, Jun 2023, Sept 2023 | Would you say that in general your health is: [Excellent, Very Good, Fair, Poor] |
| Chronic conditions diagnosed during follow up | Numeric | Sum total of ever diagnosed health conditions | Dec 2021, June 2022, December 2022, June 2023, Sept 2023 | Has a doctor, nurse, or other health professional ever told you that you had any of the following? *Select all that apply*  a) had a heart attack also called a myocardial infarction? b) have angina or coronary heart disease? c) have type 2 diabetes? d) have high blood pressure? e) have cancer? f) had asthma? g) have chronic obstructive pulmonary disease, C.O.P.D., emphysema or chronic bronchitis? h) have kidney disease (not including kidney stones, bladder infection or incontinence)? i) have HIV/AIDS? j) have immunosuppression? k) have depression? l) have post-traumatic stress disorder or PTSD? m) have chronic liver disease, including cirrhosis? n) have an anxiety disorder? o) I have not been told that I have any of the above conditions |
| Poor physical health days | Continuous | Number of days as a continuous number | July 2020, Feb 2021, May 2021, Sept 2021, Dec 2021, Mar 2022, Jun 2022, Oct 2022, Dec 2022, Apr 2023, Jun 2023, Sept 2023 | Now thinking about your physical health, which includes physical illness and injury, for how many days during the past 30 days was your physical health not good? a) _____ Number of days from 1-30 b) None c) Don’t know / Not sure |
| Poor mental health days | Continuous | Number of days as a continuous number | July 2020, Feb 2021, May 2021, Sept 2021, Dec 2021, Mar 2022, Jun 2022, Oct 2022, Dec 2022, Apr 2023, Jun 2023, Sept 2023 | Now thinking about your mental health, which includes stress, depression, and problems with emotions, for how many days during the past 30 days was your mental health not good? a) _____ Number of days from 1-30 b) None c) Don’t know / Not sure |
| Disability status | Categorical | Responses classified into three categories: [No difficulty, Some difficulty, A lot of difficulty] | July 2020, Feb 2021, May 2021, Sept 2021, Dec 2021, Mar 2022, Jun 2022, Oct 2022, Dec 2022, Apr 2023, Jun 2023, Sept 2023 | Since your last survey (ADD Qualtrics DD/Mon/YY), how much difficulty do you have engaging in daily activities (or household responsibilities) because of physical, mental, or emotional problems? a) No difficulty b) Some difficulty c) A lot of difficulty d) Don’t know / Not sure |
| Drug use | Binary | Responses collapsed to two categories: “Never”, “Once or twice”, vs. ”Daily” or “Almost daily” | Dec 2020, May 2021, Sept 2021, Dec 2021,  Mar 2022, Oct 2022, Apr 2023, Sept 2023 | In the past month how many times have you used the following: [Never, Once or twice, Weekly, Daily or almost daily]: 1) Cannabis (marijuana, pot, grass, hash, etc.), 2) Street opioids (heroin, opium, etc.), 3) Prescription opioids in a way or dose other than prescribed (fentanyl, oxycodone, hydrocodone, methadone, buprenorphine, etc.) |
| Receives government food assistance | Binary | Classify responses into two categories: 'Any government programs’ vs. ‘none’ | Feb 2021, Dec 2021, Mar 2022, Oct 2022, Apr 2023, Sept 2023 | In the past month, have you used any of the following? *Select all that apply*. [None, food pantry, soup kitchen, SNAP, pEBT, emergency food support, other] |
| Fully vaccinated for COVID-19 | Binary | Binary variable indicating whether two doses of the vaccine had been received as of the date of the survey assessment. | Feb 2021, May 2021, Sept 2021, Dec 2021, Mar 2022, Jun 2022, Oct 2022, Dec 2022, Apr 2023, Jun 2023, Sept 2023 | Have you been fully or partially vaccinated against COVID-19 with a vaccine that has received FDA approval or emergency use authorization? [Yes, No, Don’t know / Not sure]  If yes to fully or partially vaccinated in this survey: How many doses of the primary vaccine series did you receive? Primary vaccine series means either a 2-dose mRNA COVID-19 vaccine series (Moderna or Pfizer) or a single dose of Johnson & Johnson COVID-19 vaccine. If you received booster doses please do not include them here. [1, 2]  If received 1 dose only or 2 doses: When did you receive your first dose of the COVID-19 vaccine? Your vaccination card should have the date of your first shot. [Enter date: Month Day Year lookup]  If received 2 doses: When did you receive your second dose of the COVID-19 vaccine? Your vaccination card should have the date of your second shot. [Enter date: Month Day Year lookup] |
